# Physical, psychological and cognitive profile of post-COVID condition in healthcare workers, Quebec, Canada

**DOI:** 10.1101/2022.03.08.22272057

**Authors:** Sara Carazo, Danuta M. Skowronski, Robert Laforce, Denis Talbot, Emilia L. Falcone, Denis Laliberté, Geoffroy Denis, Pierre Deshaies, Sandrine Hegg-Deloye, Gaston De Serres

**Affiliations:** CHU de Québec-Laval University Research Center, Quebec City, Quebec, Canada; Biological and occupational risks unit. Institut national de santé publique du Québec, Quebec City, Quebec, Canada; Communicable Diseases and Immunization Services, BC Centre for Disease Control, Vancouver, British Columbia, Canada; Interdisciplinary Memory Clinic, Department of Neurological Sciences, CHU de Quebec; Department of Medicine, Faculty of Medicine, Laval University, Quebec City, Quebec, Canada; Social and preventive medicine department, Faculty of Medicine, Laval University, Quebec City, Quebec, Canada; Department of Medicine, Faculty of Medicine, University of Montreal, Montreal, Quebec, Canada; Center for Inflammation, Immunity and Infectious Diseases, Montreal Clinical Research Institute (IRCM), Montreal, Quebec, Canada; CIUSSS de la Capitale-Nationale, Quebec City, Quebec, Canada; CIUSSS Centre Sud de Montréal, Montreal, Quebec, Canada; McGill University, Montreal, Quebec, Canada; CISSS de Chaudière-Appalaches, Lévis, Quebec, Canada

## Abstract

**Importance:** Most adults with COVID-19 do not require hospitalization, but the subsequent risk of post-COVID condition, including associated psychological and cognitive dysfunction, remains poorly understood among non-hospitalized versus hospitalized cases.

**Objective:** To assess the prevalence and duration of post-COVID condition, including physical, psychological and cognitive symptoms.

**Design:** Case series and case-control study between December 2020 and May 2021

**Setting:** Healthcare workers in Quebec, Canada

**Participants:** Eligible cases were symptomatic healthcare workers with PCR-confirmed COVID-19 between July 2020 and May 2021. Among 17,717 contacted cases, 6061 (34%) participated. A random sample of symptomatic healthcare workers with negative PCR result between November 2020 and May 2021 served as controls. Among 11,498 contacted controls, 4390 (38%) participated.

**Exposures:** In multivariable models, sociodemographic and clinical characteristics, as well as vaccine history, were evaluated as potential risk factors. Prevalence ratios compared self-reported cognitive dysfunctions (difficulty concentrating; difficulty organizing oneself; forgetfulness; loss of necessary items) among cases with post-COVID condition to controls, adjusting for psychological distress and fatigue.

**Outcomes:** Post-COVID condition was defined by symptoms persisting ≥4 weeks or ≥12 weeks after COVID-19 onset.

**Results:** Four-week and 12-week post-COVID condition prevalences of 46% (2,746/5,943) and 40% (653/1,746), respectively, were observed among non-hospitalized cases and 76% (90/118) and 68% (27/37), respectively, among hospitalized cases. Hospitalization, female sex and age were associated with higher risk.

A substantial proportion of non-hospitalized cases with 4-week post-COVID condition often or very often reported cognitive dysfunction, including concentration (33%) or organizing (23%) difficulties, forgetfulness (20%) and loss of necessary items (10%), with no decline at 12 weeks. All four aspects of cognitive dysfunction were 2.2 to 3.0 times more prevalent among cases with post-COVID condition than in controls, but also independently associated with psychological distress and fatigue.

**Conclusions and relevance:** Post-COVID condition may be a frequent sequela of ambulatory COVID-19 in working-age adults, with important effects on cognition. With so many healthcare workers infected since the beginning of the COVID-19 pandemic, the ongoing implications for quality healthcare delivery could be profound should cognitive dysfunction and other severe post-COVID symptoms persist in a professionally-disabling way over the longer term.

**Key points:** *Question:* How common and long-lasting are the physical, psychological and cognitive effects of post-COVID condition in healthcare workers, both hospitalized and non-hospitalized?

*Findings:* The prevalence of post-COVID condition was 46% at 4 weeks and 40% at 12 weeks among non-hospitalized cases and 76% and 68% among hospitalized cases. One third of non-hospitalized healthcare workers with post-COVID condition reported cognitive impairment, which was independently associated with persistent physical symptoms, but also psychological distress and fatigue.

*Meaning:* Persistent cognitive and other professionally-disabling sequelae of COVID-19 in essential workers could have critical implications for quality healthcare delivery during and after the pandemic.

## Introduction

The COVID-19 pandemic qualifies as the most disruptive global health crisis in recent history. The persistence of symptoms long after the acute phase of the disease, as reported in a substantial proportion of cases, would add to and extend the already serious burden of this pandemic.

The Centers for Disease Control and Prevention (CDC) has defined post-COVID condition (PCC) as a wide range of physical and mental health problems present for four weeks or more after SARS-CoV-2 infection.^1^ In October 2021, to improve specificity of this heterogeneous condition and to better represent its potentially prolonged effects, the World Health Organization (WHO) defined PCC as the persistence or relapsing of symptoms at least 12 weeks after the onset of COVID-19 without alternative diagnosis.^2^

The prevalence, risk factors and duration of PCC are still poorly understood. A recent meta-analysis identified 61% and 53% of COVID-19 patients reported symptoms lasting 4-12 weeks and >12 weeks, respectively.^3^ However, hospitalized patients were over-represented and the reviewed evidence had low certainty due to self-selection bias and questionable validity of the outcome measure. Robust and valid estimates of the PCC burden are needed, particularly for non-hospitalized COVID-19 cases who comprise the vast majority of SARS-CoV-2 illness.

While PCC includes a large array of symptoms, neurocognitive symptoms, often colloquially referred to as “brain fog”, are among the most frequent and debilitating.^3,4^ Large scale epidemiological quantification of memory, concentration and executive dysfunction in PCC cases is still lacking. In particular, the contribution of PCC to cognitive dysfunctions needs to take into account the potential separate contributions of fatigue and psychological distress, also known to have deleterious effects on cognitive abilities.^5,6^

During the second and third waves of the COVID-19 pandemic in Quebec, Canada, a large case-control study was conducted to evaluate workplace exposure, infection risk and prevention in healthcare workers (HCWs), but data were also collected on the persistence of COVID-19 symptoms.^7^ In the sub-study reported here, the prevalence and risk factors for persistent COVID-19 symptoms were estimated 5 to 28 weeks after acute illness onset among predominantly non-hospitalized HCWs. PCC cases and controls were compared to assess the independent contribution of PCC, fatigue and psychological distress on persistent neurocognitive symptoms.

## Methods

### Study design and population

This case-control study included HCWs, defined as any person working in the health care sector, in Quebec, Canada. Eligible cases were HCWs with PCR-confirmed SARS-CoV-2 symptomatic infection (COVID-19) between July 12, 2020 and May 29, 2021. A random sample of HCWs who were similarly tested for COVID-19-compatible symptoms between November 14, 2020 and May 29, 2021, but who were PCR-negative and not previously infected, served as test-negative controls.

### Data collection and main variables

Contact information on SARS-CoV-2 infected HCWs and controls was obtained from the provincial COVID-19 and SARS-CoV-2 laboratory databases, which together contain all confirmed cases reported to public health and all PCR-tested individuals since the beginning of the pandemic.

HCWs were contacted by phone between December 3, 2020 and July 31, 2021. Those who agreed to participate completed a standardized 30-minute online (97%) or phone (3%) questionnaire collecting sociodemographic data, employment characteristics and COVID-19 vaccination status. Vaccination was offered to HCWs in Quebec beginning December 14, 2020.

SARS-CoV-2 infected HCWs were asked about their disease, including: symptom onset and sick leave dates; hospitalization due to COVID-19 (admission ≥24 hours); and number of weeks until full recovery. Among COVID-19 cases not completely recovered at the time of the survey, information was sought about the presence and severity (mild, moderate or severe) of 15 physical symptoms, including: fatigue, fever, shortness of breath, cough, wheezing, chest pain, headache, difficulty walking, loss of smell, loss of taste, joint or muscle pain, abdominal pain, diarrhea, sore throat and runny nose.

For all participants, the questionnaire assessed four aspects of cognitive function: 1) difficulty concentrating or maintaining attention, 2) difficulty organizing oneself, 3) forgetting things and 4) losing necessary items. Answers given on a five-level Likert scale (never, rarely, sometimes, often or very often), were considered positive if the problem occurred often or very often. Psychological distress, that may also be separately associated with cognitive dysfunction and/or PCC, was assessed with the Kessler scale or K6 (0 to 24). This scale evaluates the frequency, during the preceding month, of feeling nervous, hopeless, restless, depressed, worthless and that everything was an effort (**Supplementary Material 1**). A Kessler score of 7 to 12 indicates high psychological distress and of >12 a very high level of psychological distress.^8^

### Post-COVID condition definition

PCC was defined as the persistence of any of the 15 investigated symptoms following an acute COVID-19 disease either ≥4 weeks (PCC4w) or ≥12 weeks (PCC12w), similar to the definitions used by the CDC and WHO respectively.^1,2^ We also identified PCC4w and PCC12w with ≥1 severe symptom.

### Statistical analysis

Prevalence and binomial 95% confidence intervals (CI) were estimated. The 4-weekly prevalence trend over time for each symptom was assessed using a Cochran-Armitage test. The correlation between symptoms was explored using the Spearman correlation coefficient.

The association between PCC, socio-demographic, clinical characteristics and vaccination status was assessed estimating prevalence ratios (PR) with log-binomial regression models that included all acute COVID-19 cases.

The association between cognitive dysfunction and PCC, psychological distress and fatigue was assessed with PR using robust Poisson comparing PCC4w cases with SARS-CoV-2 test-negative controls, adjusting for sociodemographic characteristics.

The impact of PCC on the duration of sick leave (excluding students and volunteers) was assessed with mean ratios using a negative binomial regression including all cases (based on time from illness onset to survey for those still on sick leave) or only cases with finished sick leave using inverse probability weighting to account for the exclusion of those still on sick leave.

Analysis were performed using SAS version 9.4 (SAS Institute Inc).

### Ethical aspects

This study was approved by the research ethics committee of the Centre hospitalier universitaire de Québec–Université Laval. All participants gave verbal consent during the phone contact and electronic consent at the beginning of the survey.

## Results

### Population

Of the 31,091 HCWs infected with SARS-CoV-2 in Québec during the study period, 26,548 were called and 17,717 were reached (**Supplementary Figure 1**). Inclusion criteria were not met by 549 (3%), 3,485 (20%) refused to participate and 5,791 (33%) did not complete the questionnaire. Among those who answered the questionnaire, 1,831 were excluded: 140 answered the survey within 4 weeks from illness onset, 1,188 had asymptomatic infection and 503 reported symptoms lasting ≥4 weeks but had recovered at the time of the survey and were not queried about which symptom(s) persisted at least 4 weeks.

The PCC4w analysis included 6,061 HCWs with COVID-19 of which 118 (2%) had been hospitalized and 5,943 (98%) were non-hospitalized cases. The PCC12w analysis included 1,783 HCWs (37 (2%) hospitalized and 1,746 (98%) non-hospitalized). For controls, 20,770 were called and 11,498 were reached. Among them, 1,243 (11%) did not meet inclusion criteria, 2,527 (22%) refused to participate and 3,338 (29%) did not complete the survey, resulting in 4,390 controls for the analysis (**Supplementary Figure 1**).

Compared to all SARS-CoV-2 infected HCWs reported provincially, case-participants were similar for sex (78% vs 79% women), age distribution (49% vs 48% <40 years old), and percentage hospitalized (0.9% vs 1.2%).

Both hospitalized and non-hospitalized COVID-19 cases completed the questionnaire on average 10 weeks (median 9 weeks, range 5-35 weeks) after illness onset. Compared to SARS-CoV-2 infected participants, controls were younger, more often women, Caucasian and working in occupations without direct patient care (**Supplementary Table 1**).

### Prevalence and symptoms of post-COVID condition

Overall, 46.2% (95% CI: 45.0 – 47.5) of the non-hospitalized COVID-19 cases reported symptoms persisting ≥4 weeks (17.4% with at least one severe symptom) and 39.9% (95% CI: 38.3 – 41.5) had symptoms persisting ≥12 weeks. Among hospitalized COVID-19 cases, 76.3% (95% CI: 67.1 – 82.5) reported symptoms persisting ≥4 weeks (39.8% with ≥1 severe symptom) and 67.6% (95% CI: 57.7 – 79.1) had symptoms persisting ≥12-weeks (35.1% with ≥1 severe symptom) (**Table 1**).

**Table 1.** Prevalence of self-reported post-COVID condition according to different definitions for non-hospitalized and hospitalized COVID-19 cases in Quebec healthcare workers

| PCC definition: | Non-hospitalized COVID-19 HCWs |  |  |  | Hospitalized COVID-19 HCWs |  |  |  |
| --- | --- | --- | --- | --- | --- | --- | --- | --- |
|  | N | N | Prevalence |  | N | N | Prevalence |  |
|  | total | PCC | % | 95% CI<sup>a</sup> | total | PCC | % | 95% CI<sup>a</sup> |
| **≥4-week PCC** |  |  |  |  |  |  |  |  |
| a. Any symptom | 5943 | 2746 | 46.2 | 45.0 – 47.5 | 118 | 90 | 76.3 | 67.1 – 82.5 |
| b. ≥ 1 severe symptom | 5943 | 1033 | 17.4 | 16.5 – 18.4 | 118 | 47 | 39.8 | 30.8 – 48.0 |
| **≥12-week PCC** |  |  |  |  |  |  |  |  |
| c. Any symptom | 1746 | 653 | 37.4 | 35.2 – 39.7 | 37 | 27 | 73.0 | 60.0 – 88.8 |
| d. ≥ 1 severe symptom | 1746 | 245 | 14.0 | 12.5 – 15.8 | 37 | 13 | 35.1 | 22.7 – 54.4 |

<sup>a</sup> Binomial 95% confidence intervals
Abbreviations: CI, confidence interval; HCW, healthcare worker; mod, moderate; PCC, post-COVID condition

Persistent symptoms at 4-7 weeks post-COVID-19 onset were reported by half (49.6%) of non-hospitalized cases compared to 36.6% at 24-27 weeks (trend test: P<.001). The proportion with ≥1 severe symptom (18,7% to 13.6%) showed little decline with time from illness onset (trend tests: P=.93) (**Figure 1**).

**Figure 1.**
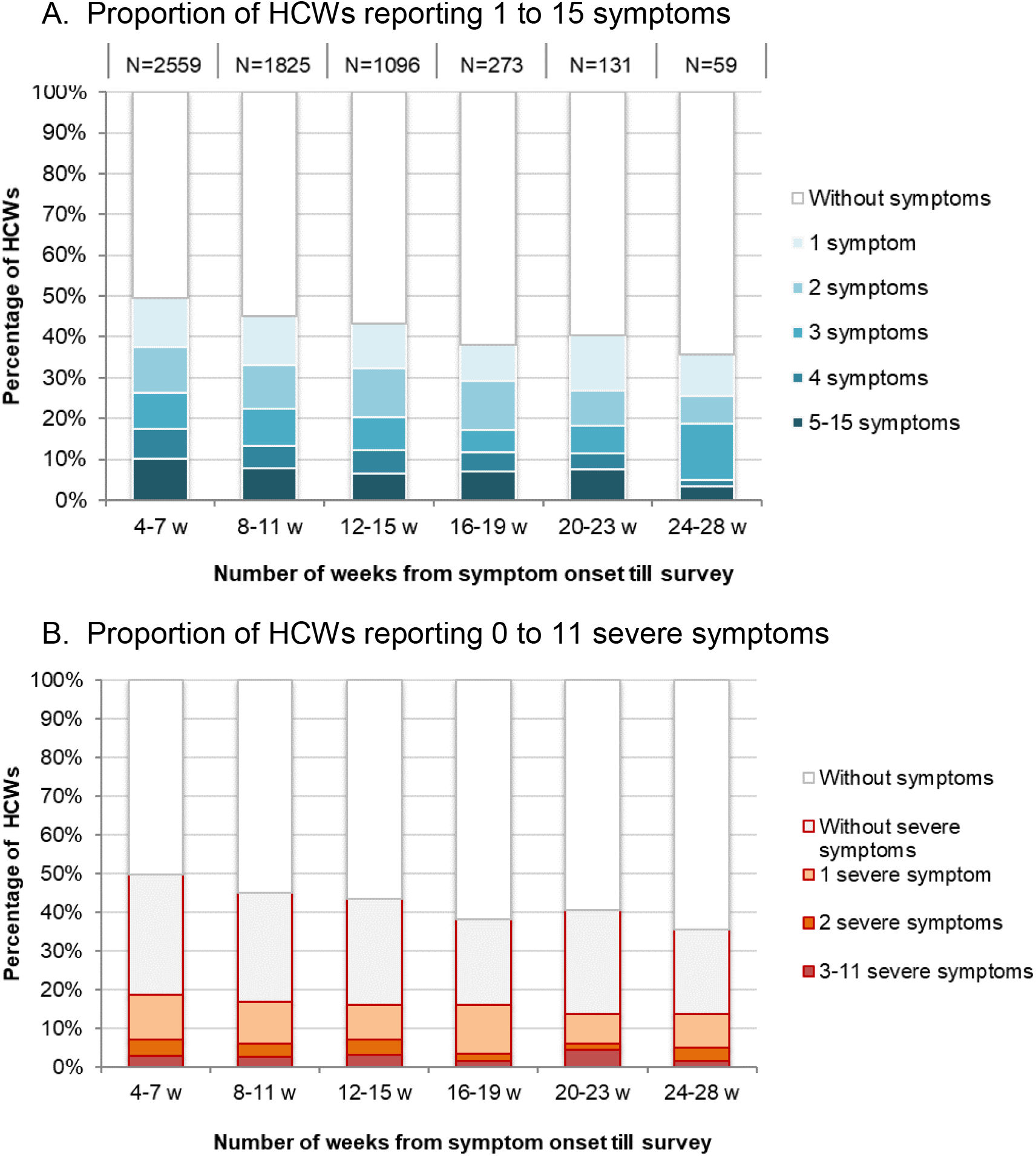
Distribution of the number of symptoms (A) and number of severe symptoms (B) reported by non-hospitalized healthcare workers, by time from COVID-19 illness onset to survey Abbreviations: HCW, healthcare worker

Among non-hospitalized and hospitalized cases, the most frequent symptoms lasting ≥4 weeks were: fatigue (30% vs 64%), loss of smell or taste (20% vs 17%), shortness of breath (20% vs 56%), cognitive dysfunction (15% vs 33%), headache (13% vs 23%), joint and muscular pain (10% vs 22%) (**Figure 2**). Symptoms more often rated as severe were loss of smell/ taste, fatigue and headache among non-hospitalized, and shortness of breath, headache and joint and muscular pain among hospitalized cases (**Figure 2**). Among cases with persistent post-COVID fatigue, 99% also reported pre-COVID fatigue; the level of fatigue however was rated as mild by 84% pre-COVID-19 and moderate (63%) or severe (28%) post-COVID-19. The overall prevalence of these symptoms slowly decreased over time, but the prevalence of severe symptoms barely changed (**Figure 3** and **Supplementary Table 2**).

**Figure 2.**
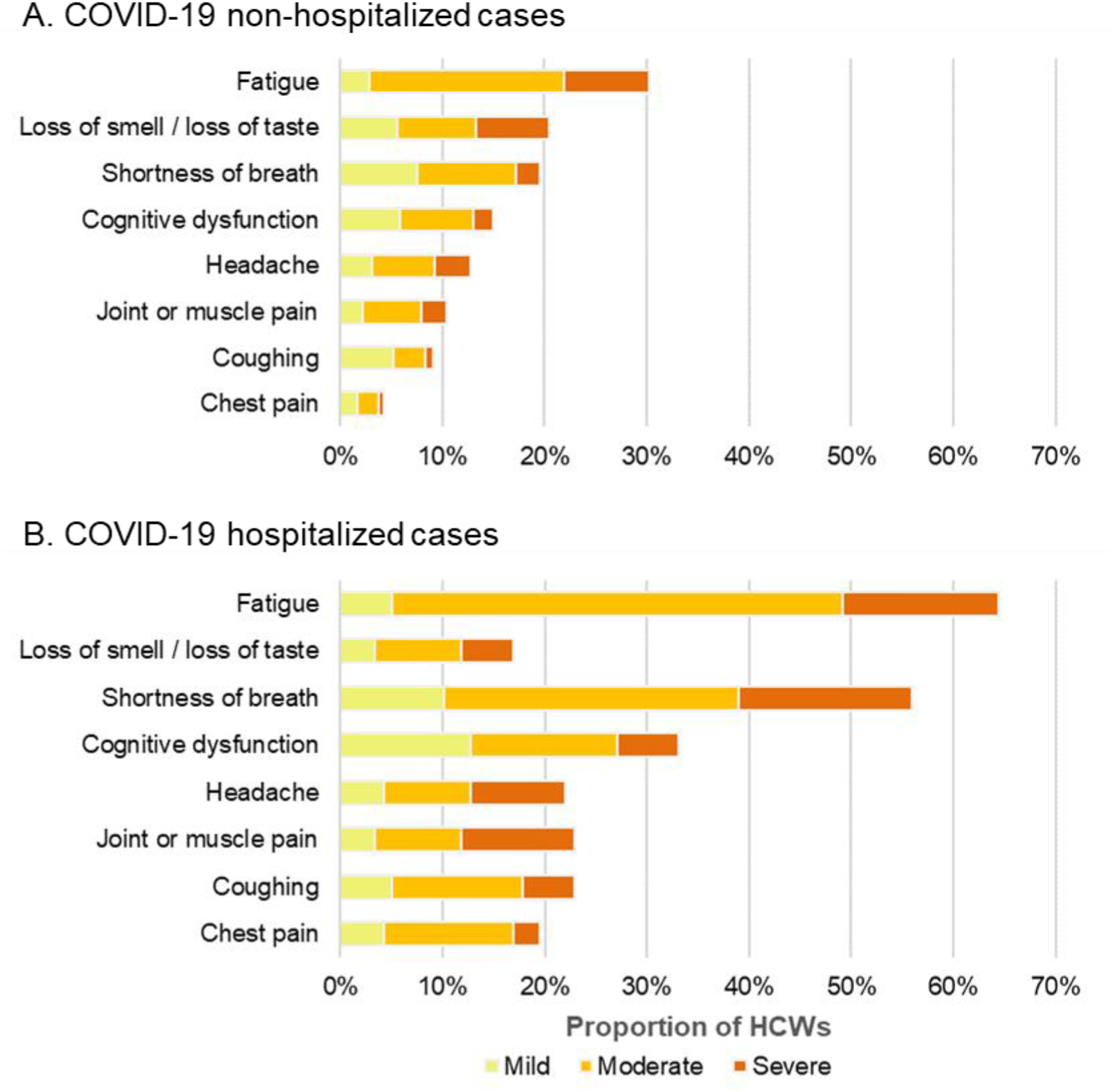
Prevalence and severity of the main symptoms still present ≥4 weeks after illness onset reported by non-hospitalized (A) and hospitalized (B) healthcare worker COVID-19 cases Note: Cognitive dysfunction defined as self-reporting often or very often presenting difficulty to concentrate or maintain attention, difficulty to organize oneself, forgetting things or loosing necessary items among those who did not present it before being infected with SARS-CoV-2

**Figure 3.**
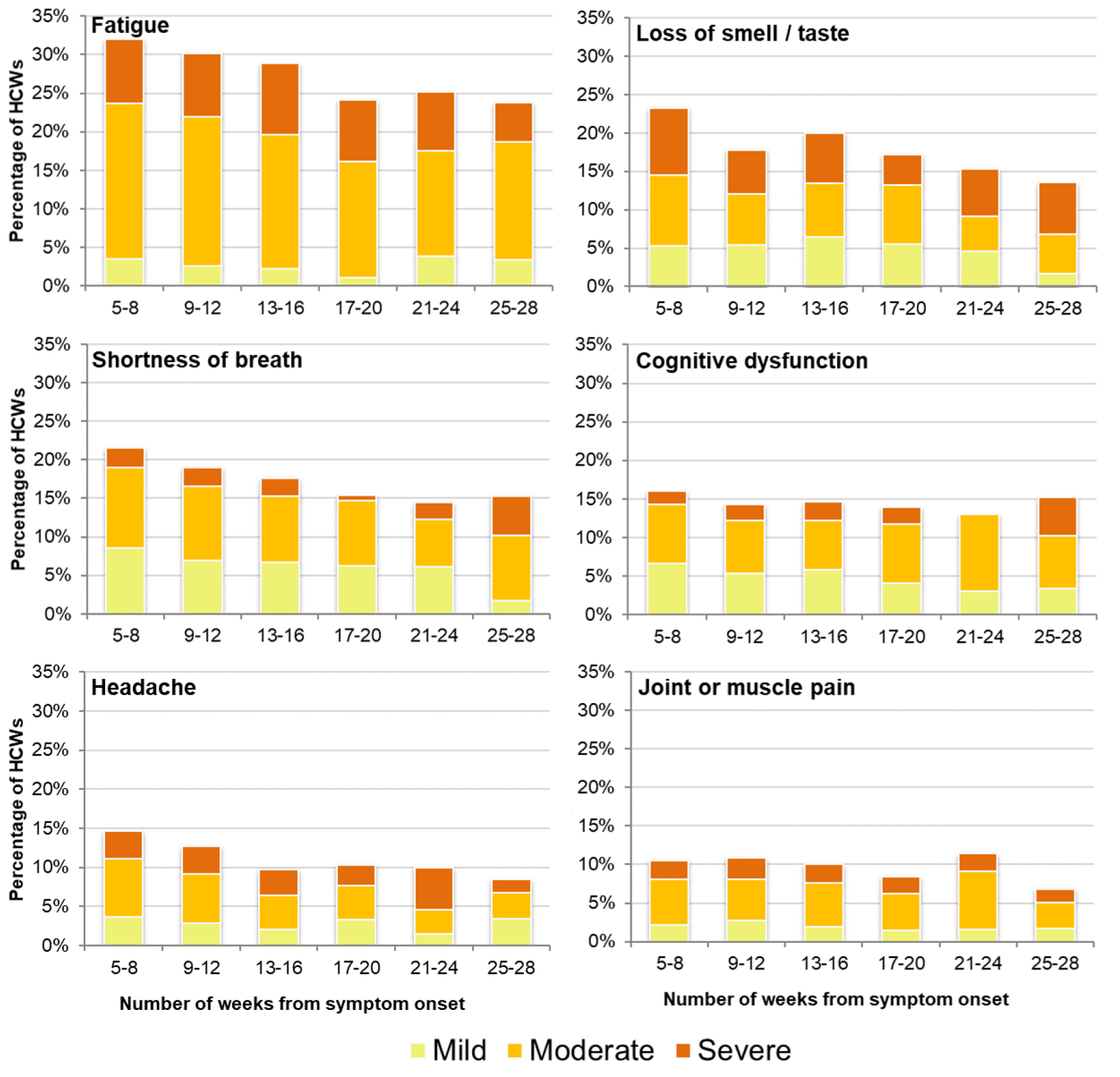
Prevalence of the 6 most commonly reported symptoms among non-hospitalized healthcare worker COVID-19 cases by time from illness onset to survey, and by severity Note: Cognitive dysfunction defined as self report of often or very often experiencing difficulty concentrating or maintaining attention, difficulty organizing oneself, forgetting things or loosing necessary items among those who did not experience these symptoms before their SARS-CoV-2 illness

The correlation between symptoms was generally greater for symptoms of the same system (**Supplementary Table 3**). Loss of smell /taste was the only symptom negatively or not correlated with any other symptom.

### Self-reported cognitive dysfunctions

The prevalence of self-reported difficulty concentrating, difficulty organizing oneself, forgetfulness and loss of necessary items often or very often was 33%, 23%, 20% and 10%, respectively, among non-hospitalized PCC4w cases, compared to 39%, 28%, 34% and 15% among hospitalized PCC4w cases (**Figure 4**). These impairments were no less frequent at 12 weeks for non-hospitalized or hospitalized cases. The report of often or very often losing necessary items, a proposed indicator of significant memory loss, was least frequent, but was significantly associated with PCC severity (6%, 26% and 68% for mild, ≥1 moderate and ≥1 severe symptom; P<.001). Those losing necessary items also experienced difficulty concentrating (84%), difficulty organizing oneself (72%) and forgetfulness (78%) (**Supplementary Table 4**).

**Figure 4.**
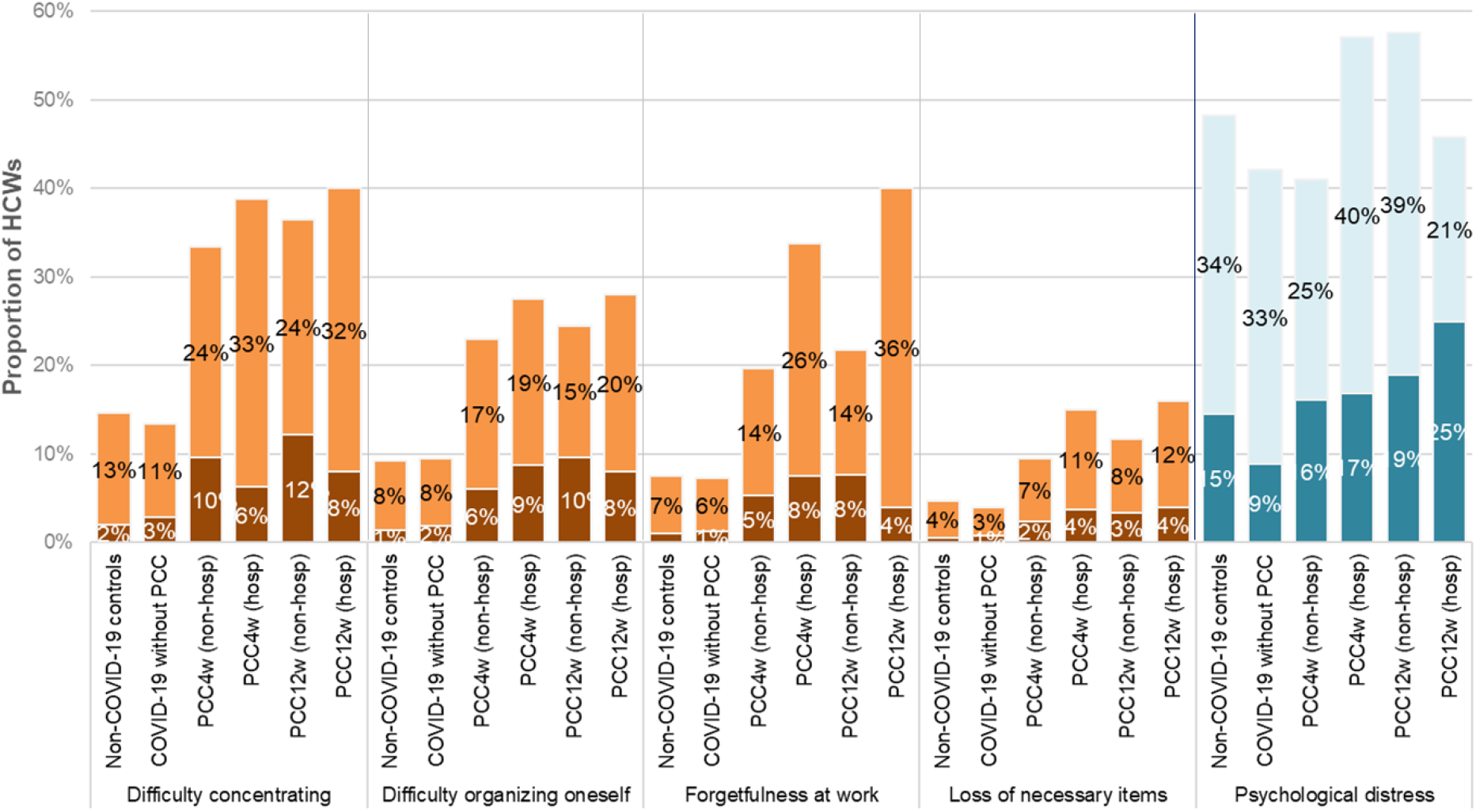
Prevalence of self-reported cognitive dysfunctions and psychological distress among non-COVID-19 controls, COVID-19 cases without post-COVID condition (PCC) and non-hospitalized and hospitalized cases with PCC, Quebec healthcare workers Abbreviations: HCW, healthcare worker; PCC4w, ≥4-week post-COVID condition; PCC12w, ≥12-week post-COVID condition

In stratified analysis, all four aspects of cognitive dysfunction were more frequent in PCC4w cases compared to controls and steadily increased with the level of psychological distress and the severity of fatigue (**Supplementary Table 5** and **Supplementary Figure 2**). In multivariable models all aspects of cognitive dysfunction were independently associated with PCC4w, psychological distress and fatigue (**Table 2**). Difficulty concentrating, difficulty organizing oneself, forgetfulness and losing necessary items were, respectively, 2.6 (95% CI: 2.4 – 2.8), 2.8 (95% CI: 2.5 – 3.1), 3.0 (95% CI: 2.6 – 3.4) and 2.2 (95% CI: 1.9 – 2.7) more common in PCC4w cases compared to controls; and 4.6 (95% CI: 4.0 – 5.3), 5.6 (95% CI: 4.6 – 6.7), 5.2 (95% CI: 4.2 – 6.4) and 4.3 (95% CI: 3.2 – 5.8) times more prevalent in individuals with very high psychological distress compared to those without distress. The four aspects of cognitive dysfunction were 2.8 (95% CI: 2.4 – 3.2), 2.9 (95% CI: 2.5 – 3.5), 3.1 (95% CI: 2.6 – 3.8) and 3.8 (95% CI: 2.8 – 5.1) times more frequent in those with severe fatigue compared to those without fatigue (**Table 2**).

**Table 2.** Prevalence of cognitive dysfunctions in cases with ≥4-week post-COVID condition and non-COVID-19 controls with or without psychological distress and their independent adjusted prevalence ratios among Quebec healthcare worker cases

| N=6962 | Cognitive dysfunction |  |  |  |  |  |  |  |  |  |  |  |
| --- | --- | --- | --- | --- | --- | --- | --- | --- | --- | --- | --- | --- |
|  | Difficulty concentrating<sup>a</sup> |  |  | Difficulty organizing oneself<sup>a</sup> |  |  | Forgetfulness<sup>a</sup> |  |  | Loss of important items<sup>a</sup> |  |  |
| Evaluated expositions: | P% | aPR<sup>b</sup> | 95% CI | P% | aPR<sup>b</sup> | 95% CI | P% | aPR<sup>b</sup> | 95% CI | P% | aPR<sup>b</sup> | 95% CI |
| **Post-COVID condition** |  |  |  |  |  |  |  |  |  |  |  |  |
| Non-COVID-19 controls | 14.6 | ref |  | 9.3 | ref |  | 7.6 | ref |  | 4.7 | ref |  |
| $\geq 4$-week PCC cases | 33.4 | **2.6** | 2.4 – 2.8 | 23.1 | **2.8** | 2.5 – 3.1 | 20.0 | **3.0** | 2.6 – 3.4 | 9.8 | **2.2** | 1.9 – 2.7 |
| **Psychological distress** |  |  |  |  |  |  |  |  |  |  |  |  |
| No (K6<7) | 8.1 | ref |  | 4.7 | ref |  | 4.1 | ref |  | 2.3 | ref |  |
| High (K6=7 to 12) | 24.1 | **2.4** | 2.1 – 2.8 | 15.9 | **2.7** | 2.3 – 3.3 | 13.7 | **2.7** | 2.2 – 3.4 | 7.3 | **2.4** | 1.8 – 3.1 |
| Very high (K6$\geq 13$) | 53.8 | **4.6** | 4.0 – 5.3 | 38.1 | **5.6** | 4.6 – 6.7 | 31.5 | **5.2** | 4.2 – 6.4 | 17.2 | **4.3** | 3.2 – 5.8 |
| **Fatigue** |  |  |  |  |  |  |  |  |  |  |  |  |
| No or mild | 10.0 | ref |  | 6.7 | ref |  | 5.7 | ref |  | 2.6 | ref |  |
| Moderate | 21.1 | **1.8** | 1.6 – 2.1 | 13.2 | **1.7** | 1.4 – 2.0 | 10.8 | **1.7** | 1.4 – 2.0 | 6.0 | **2.0** | 1.5 – 2.7 |
| Severe | 47.0 | **2.8** | 2.4 – 3.2 | 33.8 | **2.9** | 2.5 – 3.5 | 29.9 | **3.1** | 2.6 – 3.8 | 16.7 | **3.8** | 2.8 – 5.1 |
| **Sex** |  |  |  |  |  |  |  |  |  |  |  |  |
| Male | 15.8 | ref |  | 11.3 | ref |  | 9.7 | ref |  | 4.0 | ref |  |
| Female | 22.6 | **1.2** | 1.1 – 1.4 | 14.9 | **1.2** | 1.0 – 1.4 | 12.6 | **1.2** | 1.0 – 1.4 | 7.0 | **1.5** | 1.1 – 2.0 |
| **Age (years)** |  |  |  |  |  |  |  |  |  |  |  |  |
| 18-29 | 22.9 | ref |  | 15.1 | ref |  | 11.4 | ref |  | 8.0 | ref |  |
| 30-39 | 19.5 | **1.0** | 0.9 – 1.1 | 13.5 | **1.1** | 0.9 – 1.3 | 11.7 | **1.3** | 1.1 – 1.5 | 6.2 | **0.9** | 0.7 – 1.2 |
| 40-49 | 24.6 | **1.1** | 1.0 – 1.3 | 15.7 | **1.1** | 0.9 – 1.3 | 13.7 | **1.2** | 1.0 – 1.5 | 6.5 | **0.8** | 0.7 – 1.1 |
| 50-59 | 22.2 | **1.0** | 0.9 – 1.2 | 15.1 | **1.1** | 0.9 – 1.3 | 12.9 | **1.2** | 1.0 – 1.5 | 6.6 | **0.9** | 0.7 – 1.2 |
| 60-80 | 12.8 | **0.8** | 0.4 – 1.0 | 7.7 | **0.7** | 0.5 – 1.0 | 6.3 | **0.8** | 0.5 – 1.1 | 4.2 | **0.7** | 0.4 – 1.3 |

<sup>a</sup>For each manifestation: yes corresponds to a reported current frequency of often or very often
<sup>b</sup>Robust Poisson regression models adjusted for sex, age (5 categories: 18-29, 30-39, 40-49, 50-59, $>60$ years), race / ethnicity (6 categories: Caucasian, Black, Hispanic, Arab, Asiatic, other), occupation (5 categories: Physician, nurse, nurse assistant, patient healthcare assistant, housekeeping, administration /management, psychosocial workers, other), fatigue (3 categories: none or mild, moderate, severe), and psychological distress (3 categories: not $<7$ ; high= 7-12, very high $\geq 13$ in the Kessler scale).
Abbreviations: aPR, adjusted prevalence ratio; K6, Kessler scale; P, prevalence; PCC, post-COVID condition; Ref, reference category

### Risk factors for Post-COVID Condition

PCC risk was higher in hospitalized compared to non-hospitalized acute COVID-19 cases, in those aged ≥40 years and in females (**Table 3, Supplementary Table 6** and **Figure 5**). There was no specific HCW occupation at higher risk of PCC. Only 4% of cases had been vaccinated with at least one dose ≥14 days before illness onset, and their risk compared to unvaccinated cases was lower for PCC4w (PR=0.8, 95% CI: 0.7 – 0.9), but not PCC12w (PR=0.9, 95% CI: 0.7 – 1.1), although sample size was limited (**Supplementary Table 6**).

**Table 3.** Risk factors for post-COVID condition among COVID-19 healthcare workers in Quebec, according to varying case definitions (multivariable log-binomial regression model)

| N | Post-COVID condition definition |  |  |  |  |  |
| --- | --- | --- | --- | --- | --- | --- |
|  | ≥4-week PCC<br>(any symptom) |  | ≥4-week PCC-S<br>(≥1 severe symptom) |  | ≥12-week PCC<br>(any symptom) |  |
|  | 6061 |  | 6061 |  | 1783 |  |
|  | PR | 95% CI | PR | 95% CI | PR | 95% CI |
| **Age** (ref= 18-29 years) |  |  |  |  |  |  |
| 30-39 | 1.2 | 1.1 – 1.3 | 1.1 | 0.9 – 1.3 | 1.1 | 1.0 – 1.3 |
| 40-49 | 1.4 | 1.3 – 1.5 | 1.4 | 1.2 – 1.7 | 1.3 | 1.1 – 1.5 |
| 50-59 | 1.4 | 1.2 – 1.5 | 1.3 | 1.1 – 1.5 | 1.1 | 1.0 – 1.3 |
| 60-80 | 1.3 | 1.1 – 1.4 | 1.0 | 0.8 – 1.4 | 1.1 | 0.9 – 1.3 |
| **Sex** (ref=men) |  |  |  |  |  |  |
| Women | 1.2 | 1.1 – 1.3 | 1.5 | 1.3 – 1.7 | 1.1 | 1.0 – 1.2 |
| **Race / ethnicity** (ref=Caucasian) |  |  |  |  |  |  |
| Black | 0.7 | 0.6 – 0.7 | 0.7 | 0.5 – 0.9 | 0.8 | 0.6 – 0.9 |
| Hispanic | 0.8 | 0.7 – 1.0 | 1.0 | 0.7 – 1.3 | 0.9 | 0.7 – 1.1 |
| Arab | 0.7 | 0.6 – 0.9 | 0.7 | 0.5 – 1.1 | 0.9 | 0.7 – 1.1 |
| Asiatic | 0.8 | 0.6 – 0.9 | 0.7 | 0.4 – 1.1 | 0.9 | 0.7 – 1.3 |
| Other /NR | 1.0 | 0.9 – 1.1 | 1.1 | 0.9 – 1.4 | 1.0 | 0.8 – 1.2 |
| **Occupation** (ref=admin) |  |  |  |  |  |  |
| Physicians | 0.7 | 0.6 – 0.9 | 0.6 | 0.4 – 0.9 | 0.9 | 0.7 – 1.1 |
| Nurses | 1.1 | 1.0 – 1.2 | 1.1 | 0.9 – 1.3 | 0.9 | 0.8 – 1.1 |
| Nurse assistants | 1.1 | 1.0 – 1.2 | 1.3 | 1.1 – 1.7 | 1.0 | 0.8 – 1.2 |
| Patient healthcare assistants | 1.0 | 0.9 – 1.1 | 1.3 | 1.0 – 1.5 | 1.0 | 0.8 – 1.1 |
| Housekeeping | 1.0 | 0.9 – 1.2 | 1.1 | 0.8 – 1.6 | 0.9 | 0.6 – 1.2 |
| Psychosocial workers | 1.0 | 0.9 – 1.2 | 0.8 | 0.5 – 1.2 | 0.9 | 0.7 – 1.2 |
| Other | 0.9 | 0.8 – 1.0 | 1.0 | 0.8 – 1.3 | 0.8 | 0.7 – 0.9 |
| **Vaccination status**<sup>a</sup> (ref=Unvaccinated) |  |  |  |  |  |  |
| 1 dose 0-13 days | 1.0 | 0.8 – 1.1 | 1.0 | 0.7 – 1.3 | 0.9 | 0.7 – 1.1 |
| ≥1 dose ≥14 days | 0.8 | 0.7 – 0.9 | 0.8 | 0.6 – 1.1 | 0.9 | 0.7 – 1.1 |
| **Severity of acute disease**<br>(ref=non-hosp.) |  |  |  |  |  |  |
| Hospitalized | 1.5 | 1.4 – 1.7 | 2.1 | 1.7 – 2.6 | 1.6 | 1.3 – 2.1 |

<sup>a</sup> Vaccinated with 1 dose 0-13 days before illness onset or vaccinated with at least 1 dose ≥14 days before illness onset (13, 0 and 5 cases vaccinated with 2 doses for each PCC definition)
Abbreviations: admin, administration and management work; NR, no response; PCC, post-COVID condition; PCC-S, post-COVID condition with at least one severe symptom; ref, reference; PR, prevalence ratio

**Figure 5.**
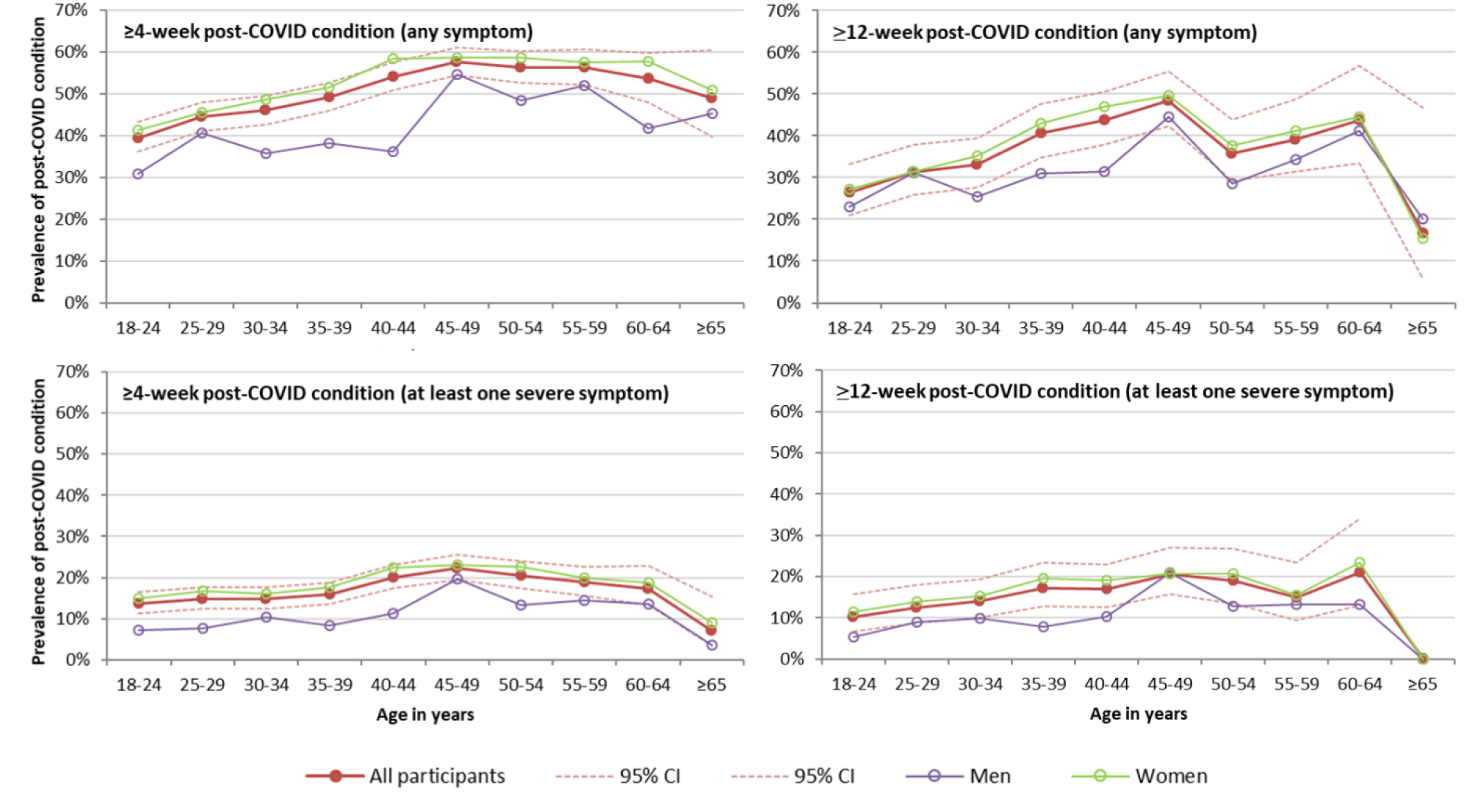
Post-COVID condition prevalence among non-hospitalized COVID-19 cases by definition, age group and sex in Quebec healthcare workers Abbreviations: CI, confidence interval

### Impact of Post-COVID condition on duration of sick leave

In a multivariable model, PCC4w cases experienced 50% longer sick leave (95% CI: 1.4 – 1.5) compared to cases whose symptoms lasted <4 weeks. Compared to non-PCC cases, a higher proportion of PCC4w cases were still on sick leave at survey (9.8% vs 1.3%) and did not feel fully recovered when returning to work (73.1% vs 26.4%) (**Supplementary Table 7**).

## Discussion

This analysis, including >6000 HCW COVID-19 cases surveyed 5-28 weeks after illness onset is, to our knowledge the largest study to date to estimate the prevalence of PCC in a representative sample of laboratory-confirmed COVID-19 adult cases, both hospitalized and non-hospitalized. COVID-19 symptoms persisting ≥4 weeks were reported by about three-quarters (76%) of hospitalized and nearly half (46%) of non-hospitalized COVID-19 cases with little decline at ≥12 weeks when about two-thirds (67.6%) and 40%, respectively, still reported persistent symptoms. Self-reported cognitive dysfunctions were highly prevalent and two to three times more frequent in PCC4w cases than controls, but were also strongly and independently associated with psychological distress and fatigue.

Similar to our results, a meta-analysis of 63 studies estimated that 61% of laboratory-confirmed COVID-19 cases had symptoms lasting 4-12 weeks and 53% had symptoms lasting more than 12 weeks.^3^ Most included studies were among hospitalized patients, had smaller sample size (between 58 and 1733 participants) and were at moderate or high risk of bias mainly due to participant selection and poor validity of outcome measures, according to Reyes et al.^3^ Other studies among non-hospitalized patients have reported highly-variable PCC prevalences, ranging from 13-44% for symptoms lasting ≥4 weeks;^9–11^ 5-35% for symptoms lasting ≥8 weeks;^9,11,12^ and 14-64% for symptoms lasting 5-8 months.^13–19^ The estimation of PCC prevalence is limited by the difficulty in obtaining a representative sample of cases with participation and attrition being highly influenced by the persistence of symptoms.^15,20^ Our large cohort is representative of all HCWs with laboratory-confirmed COVID-19 in the province of Quebec and is unlikely biased by self-selection based on symptom persistence because original recruitment was for a broader study of workplace exposure and prevention, nor post-COVID condition.

A staggering >100 physical, cognitive and psychological symptoms have been reported in PCC cases.^21^ Similar to our findings, several literature reviews found that fatigue, respiratory symptoms, cognitive and mental health issues were most frequently reported.^3,20,22–24^ Memory impairment, brain fog, poor attention or difficulty thinking have been reported by other authors in 22-88% of patients with lasting symptoms,^4,23,25,26^ but these prevalences did not take into consideration the contribution of mood disorders or fatigue. These two factors were highly associated with cognitive abnormalities in our study and in other studies.^5,6^ To accurately estimate the proportion of cognitive impairment attributable to PCC is further complicated by the fact that fatigue is one of the main features of PCC, but also of psychological distress/mood disorders. While some studies found that COVID-19 patients frequently reported anxiety or depression,^20,22,27^ a systematic review including 33 studies found similar rates of anxiety or depression in the general population.^28^ In our study, very high psychological distress, which is strongly associated with anxiety and depression,^29,30^ was similarly common among PCC cases and controls and likely related to other work-related psychosocial risks and stressors afflicting HCWs during the pandemic.^31^ The prognosis of these cognitive dysfunctions is unknown but the lack of decline in prevalence over 4-28-weeks is worrisome. If persisting over the long-term, this not infrequent sequela of COVID-19 could become both personally and professionally impactful on a significant scale among the highly infected population of HCWs. Our study also underscores the significant impact of fatigue and psychological distress during the pandemic on HCW cognitive function independent of PCC status, which also has important implications for quality healthcare delivery.

As reported elsewhere, our hospitalized cases had a higher prevalence and severity of PCC than ambulatory cases,^12,14,20^ with female sex and increasing age also associated with higher PCC risk.^9,12,15,16,25,32–34^ We could not properly assess the impact of vaccination as few of our participants had been vaccinated at the time of their COVID-19. Lasting post-COVID-19 symptoms disrupt work life with 8% to 73% indicating they were unable to return to previous full-time work 3 to 8 months after acute COVID-19 ^4,11,35^. Our PCC cases not only had a longer duration of sick leave, but three quarters reported having returned to work when not yet fully recovered, also concerning with respect to quality of care.

As may be expected in the evaluation of a novel and heterogeneous condition, our study has several limitations. Using CDC and WHO case definitions for PCC based on 4-week or 12-week duration may have overestimated the prevalence of clinically-significant PCC. Clinical significance may be improved by requiring longer duration, but remains suboptimal in allowing any symptom of any intensity. Temporal patterns were identified using a single-time survey completed at different moments since illness onset and asking only about ongoing symptoms. This may have underestimated the prevalence of symptoms for shorter intervals but avoided recall bias and the selective attrition of less symptomatic patients in follow-up studies.^3,15^ We did not collect information on some potentially relevant risk factors, such as comorbidity or initial COVID-19 symptoms and their severity.^14,15,25,32,34^ Our questionnaire, designed in the fall 2020, did not collect data on insomnia or post-exertional malaise later also considered to be PCC-qualifying symptoms.^21^ Cognitive functions were not formally measured with the validated instruments neurologists use to investigate patients, and self-reporting might have over-estimated their prevalence. Physical symptoms among non-COVID-19 controls were not collected. Comparing the prevalence but also the severity of symptoms among cases and controls would have allowed better characterization of PCC and should be considered in future research. Our participants were mostly unvaccinated working adults infected by the original SARS-CoV-2 virus: findings may not be generalizable to children, elderly adults, vaccinated individuals or those infected by subsequent variants of concern. Despite these limitations, this study shows the high burden of COVID-19 sequelae among non-hospitalized adults.

In conclusion, PCC may be a frequent sequela of ambulatory COVID-19 in working-age adults, with important effects on cognition. With so many HCWs infected since the beginning of the COVID-19 pandemic, the ongoing implications for quality healthcare delivery could be profound should cognitive dysfunction and other severe PCC symptoms persist in a professionally-disabling way over the longer term.

## Data Availability

Only data contained in the manuscript is available

## Acknowledgments

We would like to thank Jasmin Villeneuve, Richard Martin (Institut national de santé du Québec), Armelle Lorcy (CHU de Québec-Université Laval Research Center), Francine Ducharme (Université de Montréal) and Bianka Paquet-Bolduc (Institut Universitaire en cardiologie et pneumologie de Québec) for their contribution to the large study conducted to evaluate COVID-19 workplace exposure, infection risk and prevention in healthcare workers in Quebec. We also thank all the healthcare workers that participated to this study.

