## Supplemental material for "Physical, psychological and cognitive profile of post-COVID condition in healthcare workers, Quebec, Canada"

### Supplementary Material

**Supplementary Figure 1.** Flowchart of study population

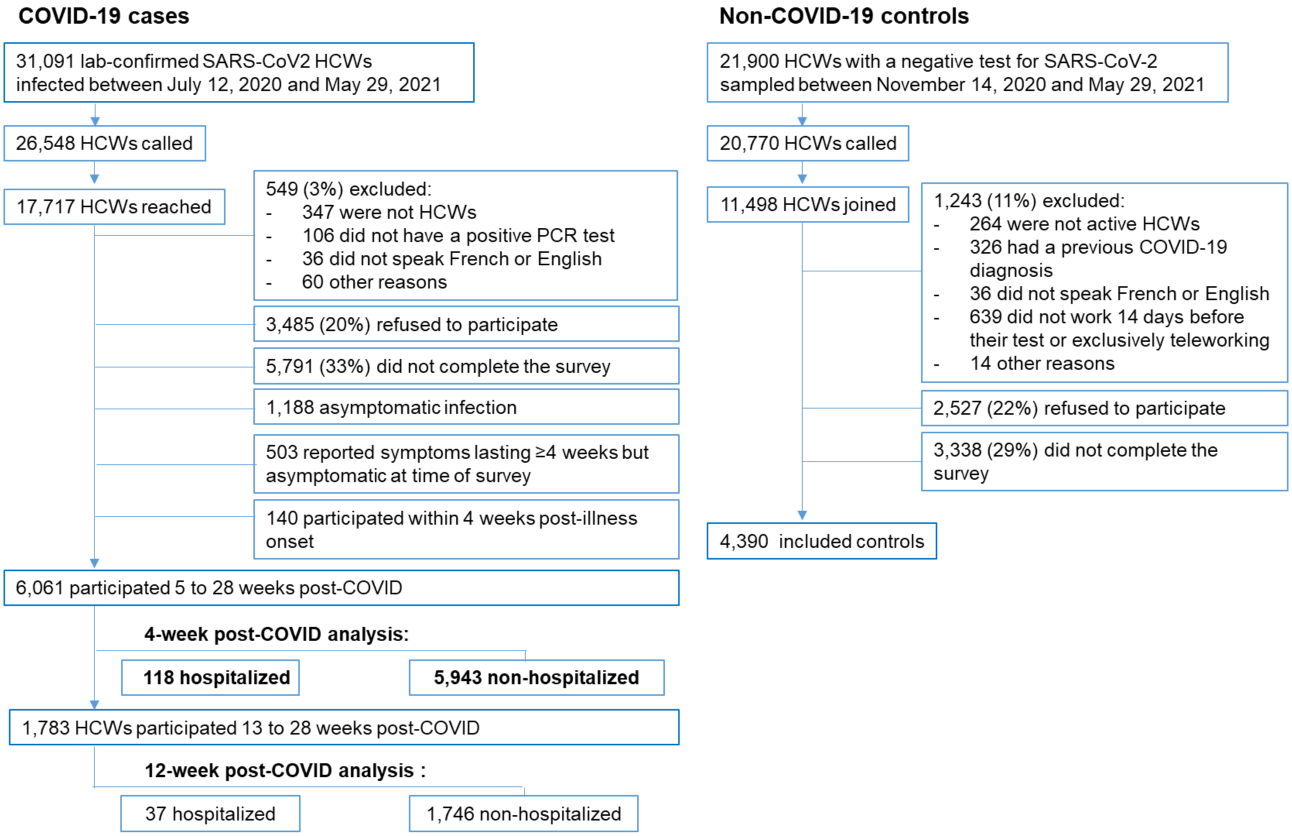

Abbreviations: HCW, healthcare worker

**Supplementary Figure 2.** Prevalence of often or very often self-reporting difficulty concentrating, difficulty organizing oneself, forgetfulness and loss of necessary items by level of fatigue and level of psychological distress (measured by Kessler scale) among cases with ≥4-week post-COVID condition and non-COVID-19 controls, Quebec healthcare workers

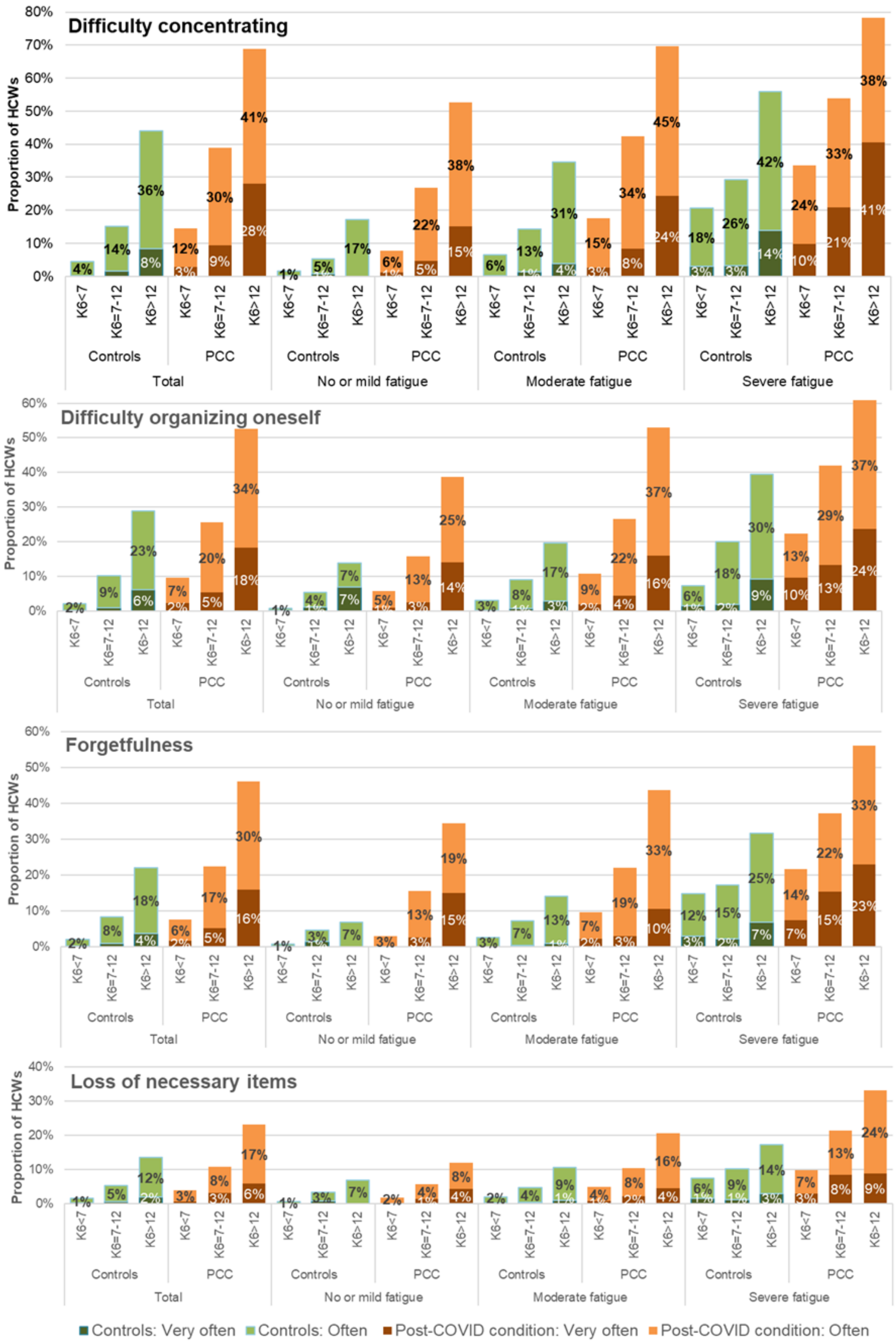

Note: Psychological distress according to the Kessler K6 questionnaire: high for K6=7-12 and very high for K6>12;
Abbreviations: HCW, healthcare worker; PCC, post-COVID condition

**Supplementary Table 1.** Characteristics of participants and comparison groups, Quebec healthcare workers

| | Type of participants | | |
|---|---|---|---|
| | Symptomatic hospitalized Covid-19 HCWs | Symptomatic non-hospitalized Covid-19 HCWs | Non-Covid-19 HCWs (controls) |
| **CHARACTERISTICS** | N (%) | N (%) | N (%) |
| Total | 118 | 5934 | 4390 |
| **Weeks from illness onset or testing to study participation**: Mean ± STD | 10.3 ± 4.5 | 10.2 ± 4.3 | 9.9 ± 3.5 |
| **Age** (years): Mean ± STD | 46.7 ± 11.9 | 40.0 ± 12.1 | 39.0 ± 10.4 |
| 18-29 | 12 (10.2) | 1410 (23.7) | 807 (18.4) |
| 30-39 | 21 (17.8) | 1547 (26.0) | 1750 (39.9) |
| 40-49 | 30 (25.4) | 1550 (26.1) | 1045 (23.8) |
| 50-59 | 39 (33.1) | 1102 (18.5) | 608 (13.9) |
| 60-80 | 16 (13.6) | 334 (5.6) | 180 (4.1) |
| **Sex**: Women | 83 (70.3) | 4704 (79.2) | 3819 (87.0) |
| **Race/ethnicity** | | | |
| White | 81 (68.6) | 4713 (79.3) | 3995 (91.0) |
| Black | 8 (6.8) | 482 (8.1) | 103 (2.4) |
| Hispanic | 10 (8.5) | 163 (2.7) | 68 (1.6) |
| Arab | 8 (6.8) | 176 (3.0) | 78 (1.8) |
| Asiatic | 2 (1.7) | 141 (2.4) | 56 (1.3) |
| Other/NR | 9 (7.6) | 268 (4.5) | 90 (2.1) |
| **Occupation** | | | |
| Physicians | 2 (1.7) | 241 (4.1) | 221 (5.0) |
| Nurses | 20 (17.0) | 1118 (18.8) | 900 (20.5) |
| Nurse assistants | 15 (12.7) | 469 (7.9) | 222 (5.1) |
| Patient healthcare assistants | 40 (33.9) | 1526 (25.7) | 523 (11.9) |
| Housekeeping | 5 (4.2) | 197 (3.3) | 40 (0.9) |
| Administration/manager | 13 (11.0) | 591 (9.9) | 618 (14.1) |
| Psychosocial worker | 5 (4.2) | 197 (3.3) | 397 (9.0) |
| Others | 18 (15.3) | 1585 (26.7) | 1469 (33.5) |
| **Vaccination status** | | | |
| Non-vaccinated | 114 (96.6) | 5484 (92.3) | 2980 (67.9) |
| 1 dose 0-13 days before | 2 (1.7) | 230 (3.9) | 280 (6.4) |
| 1 dose ≥14 days before | 2 (1.7) | 216 (3.6) | 926 (21.1) |
| 2 doses | 0 (0.0) | 13 (0.2) | 204 (4.7) |

Abbreviations: STD, standard deviation

**Supplementary Table 2.** Description of reported symptoms by severity and time since illness onset among healthcare workers in Quebec with non-hospitalized and hospitalized COVID-19, still symptomatic at the time of the survey

| | Presence | | Severity (L %) | | | Weeks from symptoms onset (C %) | | | | | |
|---|---|---|---|---|---|---|---|---|---|---|---|
| | N | % | Mild | Mod | Sev | 5-8 | 9-12 | 13-16 | 17-20 | 21-29 | P[a] |
| **NON-HOSPITALIZED** | **2746** | | | | | **1270** | **823** | **475** | **104** | **74** | |
| Fatigue | 1800 | 65.6 | 9.4 | 62.8 | 27.9 | 64.6 | 66.8 | 66.7 | 63.5 | 63.5 | 0.93 |
| Fatigue before COVID-19 | 1776 | 64.1 | 83.8 | 13.3 | 2.9 | | | | | | |
| Loss of smell / loss of taste | 1213 | 44.2 | 27.0 | 38.2 | 34.8 | 46.8 | 39.5 | 46.1 | 45.2 | 37.8 | 0.14 |
| Shortness of breath | 1160 | 42.2 | 38.5 | 49.4 | 12.2 | 43.3 | 42.2 | 40.6 | 40.4 | 37.8 | 0.16 |
| Cognitive dysfunction[b] | 894 | 35.1 | 38.7 | 48.1 | 13.2 | 34.4 | 34.3 | 37.0 | 40.0 | 37.1 | 0.06 |
| Headache | 759 | 27.6 | 24.0 | 48.8 | 27.3 | 29.5 | 28.1 | 22.5 | 26.9 | 24.3 | **<.01** |
| Joint or muscle pain | 621 | 22.6 | 21.4 | 54.4 | 24.2 | 21.3 | 24.2 | 23.2 | 22.1 | 25.7 | 0.19 |
| Cough | 542 | 19.8 | 56.6 | 35.2 | 8.1 | 24.7 | 16.3 | 15.8 | 12.5 | 9.5 | **<.01** |
| Chest pain | 254 | 9.2 | 38.2 | 50.8 | 11.0 | 9.7 | 8.6 | 8.6 | 11.5 | 9.5 | 0.86 |
| Runny nose | 232 | 8.5 | 40.5 | 49.6 | 9.9 | 9.6 | 7.8 | 5.5 | 14.4 | 6.8 | 0.22 |
| Sore throat | 178 | 6.5 | 52.8 | 37.1 | 10.1 | 8.2 | 4.7 | 5.3 | 3.9 | 8.1 | **0.02** |
| Wheezing | 159 | 5.8 | 42.1 | 47.2 | 10.7 | 5.8 | 5.2 | 7.2 | 3.9 | 5.4 | 0.28 |
| Diarrhea | 111 | 4.0 | 31.5 | 55.0 | 13.5 | 4.9 | 3.9 | 2.5 | 1.0 | 5.4 | 0.07 |
| Difficulty walking | 99 | 3.6 | 19.2 | 64.7 | 16.2 | 2.7 | 4.7 | 4.0 | 3.9 | 4.1 | 0.27 |
| Abdominal pain | 94 | 3.4 | 31.9 | 51.1 | 17.0 | 3.6 | 4.0 | 1.7 | 3.9 | 4.1 | 0.25 |
| Fever | 70 | 2.6 | 27.1 | 62.9 | 10.0 | 2.7 | 2.3 | 2.7 | 1.0 | 4.1 | 0.80 |
| **HOSPITALIZED** | **90** | | | | | **40** | **23** | **16** | **11**[c] | | |
| Fatigue | 76 | 84.4 | 7.9 | 68.4 | 23.7 | 85.0 | 87.0 | 87.5 | 72.7 | | 0.52 |
| Fatigue before COVID-19 | 75 | 83.3 | 93.3 | 2.7 | 4.0 | | | | | | |
| Loss of smell / loss of taste | 20 | 22.2 | 20.0 | 50.0 | 30.0 | 17.5 | 17.4 | 43.8 | 18.2 | | 0.28 |
| Shortness of breath | 66 | 73.3 | 18.2 | 51.5 | 30.3 | 72.5 | 87.0 | 50.0 | 81.8 | | 0.73 |
| Cognitive dysfunction[1] | 39 | 47.6 | 38.5 | 43.6 | 18.0 | 48.6 | 45.5 | 26.7 | 80.0 | | 0.53 |
| Headache | 27 | 30.0 | 14.8 | 37.0 | 48.2 | 30.0 | 34.8 | 25.0 | 27.3 | | 0.76 |
| Joint or muscle pain | 26 | 28.9 | 19.2 | 38.5 | 42.3 | 17.5 | 30.4 | 37.5 | 54.6 | | **0.01** |
| Cough | 27 | 30.0 | 22.2 | 55.6 | 22.2 | 35.0 | 34.8 | 25.0 | 9.1 | | 0.11 |
| Chest pain | 23 | 25.6 | 21.7 | 65.2 | 13.0 | 32.5 | 26.1 | 18.8 | 9.1 | | 0.09 |
| Runny nose | 8 | 8.9 | 50.0 | 25.0 | 25.0 | 10.0 | 8.7 | 6.3 | 9.1 | | 0.77 |
| Sore throat | 12 | 13.3 | 41.7 | 50.0 | 8.3 | 12.5 | 13.0 | 18.8 | 9.1 | | 0.94 |
| Wheezing | 9 | 10.0 | 22.2 | 44.4 | 33.3 | 15.0 | 4.4 | 0 | 18.2 | | 0.55 |
| Diarrhea | 6 | 6.7 | 33.3 | 16.7 | 50.0 | 5.0 | 8.7 | 6.3 | 9.1 | | 0.65 |
| Difficulty walking | 18 | 20.0 | 16.7 | 61.1 | 22.2 | 25.0 | 13.0 | 12.5 | 27.3 | | 0.69 |
| Abdominal pain | 5 | 5.6 | 20.0 | 0 | 80.0 | 7.5 | 4.4 | 0 | 9.1 | | 0.60 |
| Fever | 7 | 7.8 | 0 | 71.4 | 28.6 | 15.5 | 4.4 | 0 | 9.1 | | 0.29 |

[a] P-value for the Cochran-Armitage trend test
[b] Cognitive dysfunction defined as self-reporting often or very often presenting difficulty to concentrate or maintain attention, difficulty to organize oneself, forgetting things or loosing necessary items among those who did not present it before being infected with SARS-CoV-2
[c] 17-24 weeks (8 HCWs participated 17-20 weeks and 3 HCWs 21-24 weeks post-illness onset)
Abbreviations: C %, column percentage; HCW, healthcare worker; L %, line percentage; Mod, moderate; Sev, severe

**Supplementary Table 3.** Spearman correlation coefficients between symptoms of cases with ≥4-week post-COVID condition

| | Fatigue | Fever | Joint or muscle pain | Shortness of breath | Cough | Wheezing | Chest pain | Cognitive dysfunction | Headache | Difficulty walking | Loss of smell and/or taste | Abdominal pain | Diarrhea | Sore throat | Runny nose |
|---|---|---|---|---|---|---|---|---|---|---|---|---|---|---|---|
| **Fatigue** | 1.00 | 0.12 | 0.27 | 0.20 | 0.02 | 0.08 | 0.12 | 0.27 | 0.25 | 0.12 | -0.23 | 0.11 | 0.08 | 0.08 | 0.08 |
| p | | <.01 | <.01 | <.01 | 0.23 | <.01 | <.01 | <.01 | <.01 | <.01 | <.01 | <.01 | <.01 | <.01 | <.01 |
| **Fever** | 0.12 | 1.00 | 0.20 | 0.08 | 0.11 | 0.08 | 0.14 | 0.07 | 0.13 | 0.19 | 0.02 | 0.07 | 0.12 | 0.13 | 0.11 |
| p | | | <.01 | <.01 | <.01 | <.01 | <.01 | <.01 | <.01 | <.01 | 0.32 | <.01 | <.01 | <.01 | <.01 |
| **Joint or muscle pain** | 0.27 | 0.20 | 1.00 | 0.14 | 0.04 | 0.09 | 0.17 | 0.18 | 0.21 | 0.25 | -0.10 | 0.18 | 0.16 | 0.13 | 0.10 |
| p | | <.01 | | <.01 | 0.02 | <.01 | <.01 | <.01 | <.01 | <.01 | <.01 | <.01 | <.01 | <.01 | <.01 |
| **Shortness of breath** | 0.20 | 0.08 | 0.14 | 1.00 | 0.11 | 0.19 | 0.21 | 0.17 | 0.11 | 0.14 | -0.16 | 0.05 | 0.09 | 0.05 | 0.06 |
| p | | <.01 | <.01 | | <.01 | <.01 | <.01 | <.01 | <.01 | <.01 | <.01 | <.01 | <.01 | 0.01 | <.01 |
| **Cough** | 0.02 | 0.11 | 0.04 | 0.11 | 1.00 | 0.09 | 0.10 | 0.05 | 0.03 | 0.02 | -0.07 | 0.07 | 0.05 | 0.17 | 0.13 |
| p | | <.01 | 0.02 | <.01 | | <.01 | <.01 | 0.02 | 0.12 | 0.23 | <.01 | <.01 | 0.01 | <.01 | <.01 |
| **Wheezing** | 0.08 | 0.08 | 0.09 | 0.19 | 0.09 | 1.00 | 0.12 | 0.09 | 0.06 | 0.09 | -0.03 | 0.04 | 0.06 | 0.06 | 0.09 |
| p | | <.01 | <.01 | <.01 | <.01 | | <.01 | <.01 | <.01 | <.01 | 0.13 | 0.04 | <.01 | <.01 | <.01 |
| **Chest pain** | 0.12 | 0.14 | 0.17 | 0.21 | 0.10 | 0.12 | 1.00 | 0.10 | 0.14 | 0.16 | -0.05 | 0.15 | 0.08 | 0.12 | 0.09 |
| p | | <.01 | <.01 | <.01 | <.01 | <.01 | | <.01 | <.01 | <.01 | 0.01 | <.01 | <.01 | <.01 | <.01 |
| **Cognitive dysfunction** | 0.27 | 0.07 | 0.18 | 0.17 | 0.05 | 0.09 | 0.10 | 1.00 | 0.16 | 0.11 | -0.04 | 0.12 | 0.06 | 0.07 | 0.06 |
| p | | <.01 | <.01 | <.01 | 0.02 | <.01 | <.01 | | <.01 | <.01 | 0.04 | <.01 | <.01 | <.01 | <.01 |
| **Headache** | 0.25 | 0.13 | 0.21 | 0.11 | 0.03 | 0.06 | 0.14 | 0.16 | 1.00 | 0.08 | -0.12 | 0.15 | 0.15 | 0.10 | 0.12 |
| p | | <.01 | <.01 | <.01 | 0.12 | <.01 | <.01 | <.01 | | <.01 | <.01 | <.01 | <.01 | <.01 | <.01 |
| **Difficulty walking** | 0.12 | 0.19 | 0.25 | 0.14 | 0.02 | 0.09 | 0.16 | 0.11 | 0.08 | 1.00 | -0.01 | 0.15 | 0.15 | 0.06 | 0.06 |
| p | | <.01 | <.01 | <.01 | 0.23 | <.01 | <.01 | <.01 | <.01 | | 0.79 | <.01 | <.01 | <.01 | <.01 |
| **Loss of smell and/or taste** | -0.23 | 0.02 | -0.10 | -0.16 | -0.07 | -0.03 | -0.05 | -0.04 | -0.12 | -0.01 | 1.00 | 0.00 | 0.01 | -0.01 | -0.01 |
| p | | 0.32 | <.01 | <.01 | <.01 | 0.13 | 0.01 | 0.04 | <.01 | 0.79 | | 0.92 | 0.78 | 0.58 | 0.45 |
| **Abdominal pain** | 0.11 | 0.07 | 0.18 | 0.05 | 0.07 | 0.04 | 0.15 | 0.12 | 0.15 | 0.15 | 0.00 | 1.00 | 0.26 | 0.15 | 0.10 |
| p | | <.01 | <.01 | <.01 | <.01 | 0.04 | <.01 | <.01 | <.01 | <.01 | 0.92 | | <.01 | <.01 | <.01 |
| **Diarrhea** | 0.08 | 0.12 | 0.16 | 0.09 | 0.05 | 0.06 | 0.08 | 0.06 | 0.15 | 0.15 | 0.01 | 0.26 | 1.00 | 0.13 | 0.10 |
| p | | <.01 | <.01 | <.01 | 0.01 | <.01 | <.01 | <.01 | <.01 | <.01 | 0.78 | <.01 | | <.01 | <.01 |
| **Sore throat** | 0.08 | 0.13 | 0.13 | 0.05 | 0.17 | 0.06 | 0.12 | 0.07 | 0.10 | 0.06 | -0.01 | 0.15 | 0.13 | 1.00 | 0.21 |
| p | | <.01 | <.01 | 0.01 | <.01 | 0.00 | <.01 | 0.00 | <.01 | 0.00 | 0.58 | <.01 | <.01 | | <.01 |
| **Runny nose** | 0.08 | 0.11 | 0.10 | 0.06 | 0.13 | 0.09 | 0.09 | 0.06 | 0.12 | 0.06 | -0.01 | 0.10 | 0.10 | 0.21 | 1.00 |
| p | | <.01 | <.01 | 0.00 | <.01 | <.01 | <.01 | 0.00 | <.01 | 0.00 | 0.45 | <.01 | <.01 | <.01 | |

Note: Green case = correlation >0.15, Orange case = no or negative correlation

**Supplementary Table 4.** Severity of post-COVID condition (PCC) and prevalence of physical symptoms, psychological distress and self-reported cognitive dysfunctions (difficulty concentrating, difficulty organizing oneself and forgetfulness) among PCC cases in Quebec healthcare workers reporting often or very often losing important items

| | | Loss of necessary items | | |
|---|---|---|---|---|
| | | Often / very often | Never / rarely / sometimes | |
| | | N (%) | N (%) | p |
| | | 257 | 2384 | |
| **Severity of PCC** | Only mild symptom | 14 (5.6) | 450 (18.9) | <.01 |
| | ≥ 1 moderate symptom | 66 (25.7) | 1134 (47.6) | |
| | ≥ 1 severe symptom | 177 (68.8) | 800 (33.6) | |
| **Number of symptoms** | 1 | 12 (4.7) | 619 (26.0) | <.01 |
| | 2 | 30 (11.7) | 598 (25.1) | |
| | 3 | 55 (21.4) | 453 (19.0) | |
| | 4 | 41 (16.0) | 315 (13.2) | |
| | 5 to 15 | 119 (46.3) | 399 (16.7) | |
| **Number of severe symptoms** | 0 | 74 (28.8) | 1566 (65.7) | <.01 |
| | 1 | 74 (28.8) | 542 (22.7) | |
| | 2 | 41 (16.0) | 169 (7.1) | |
| | 3 to 11 | 68 (26.5) | 107 (4.5) | |
| **Psychological distress** | K6<7 | 43 (16.7) | 1081 (45.3) | <.01 |
| | K6=7-12 | 117 (45.5) | 978 (41.0) | |
| | K6>12 | 97 (37.7) | 325 (13.6) | |
| **Other cognitive dysfunctions** | Difficulty concentrating | 217 (84.4) | 664 (27.9) | <.01 |
| | Difficulty organizing oneself | 186 (72.4) | 424 (17.8) | <.01 |
| | Forgetfulness | 200 (77.8) | 326 (13.7) | <.01 |
| **Physical symptoms** | Fatigue | 220 (85.6) | 1533 (64.3) | <.01 |
| | Shortness of breath | 152 (59.1) | 998 (41.9) | <.01 |
| | Headache | 111 (43.2) | 624 (26.2) | <.01 |
| | Joint or muscle pain | 110 (42.8) | 499 (20.9) | <.01 |
| | Loss of smell / loss of taste | 108 (42.0) | 1034 (43.4) | 0.68 |
| | Cough | 59 (23.0) | 487 (20.4) | 0.34 |
| | Chest pain | 42 (16.3) | 220 (9.2) | <.01 |
| | Runny nose | 40 (15.6) | 196 (8.2) | <.01 |
| | Sore throat | 34 (13.2) | 146 (6.1) | <.01 |
| | Difficulty walking | 30 (11.7) | 81 (3.4) | <.01 |
| | Wheezing | 27 (10.5) | 129 (5.4) | <.01 |
| | Abdominal pain | 26 (10.1) | 68 (2.9) | <.01 |
| | Diarrhea | 19 (7.4) | 92 (3.9) | <.01 |
| | Fever | 17 (6.6) | 52 (2.2) | <.01 |

Abbreviations: PCC, post-COVID condition

**Supplementary Table 5.** Prevalence of cognitive symptoms, fatigue and psychological distress among cases with post-COVID condition and non-COVID-19 controls, Quebec healthcare workers

| | PCC (hospitalized) | PCC (non-hospitalized) | Non-COVID-19 controls | |
|---|---|---|---|---|
| | N (%) | N (%) | N (%) | p |
| | 77 | 2498 | 4390 | |
| **Cognitive dysfunctions** | | | | |
| Difficulty concentrating | | | | <.01 |
| Often | 24 (31.2) | 591 (23.7) | 553 (12.6) | |
| Very often | 5 (6.5) | 241 (9.7) | 87 (2.0) | |
| Difficulty organizing oneself | | | | <.01 |
| Often | 13 (16.9) | 423 (16.9) | 346 (7.9) | |
| Very often | 7 (9.1) | 151 (6.0) | 60 (1.4) | |
| Forgetfulness | | | | <.01 |
| Often | 20 (26.0) | 355 (14.2) | 288 (6.6) | |
| Very often | 6 (7.8) | 133 (5.3) | 43 (1.0) | |
| Loss of necessary items | | | | <.01 |
| Often | 8 (23.4) | 179 (7.2) | 186 (4.2) | |
| Very often | 3 (3.9) | 61 (2.4) | 22 (0.5) | |
| **Psychological distress** | | | | 0.23 |
| No (K6<7) | 33 (42.9) | 1057 (42.3) | 1972 (44.9) | |
| High (K6=7-12) | 31 (40.3) | 1039 (41.6) | 1779 (40.5) | |
| Very high (K6>12) | 33 (42.9) | 1057 (41.3) | 1972 (44.9) | |
| **Fatigue** | | | | <.01 |
| No or mild | 16 (20.8) | 1016 (40.7) | 1283 (29.2) | |
| Moderate | 46 (59.7) | 1029 (41.2) | 2461 (56.0) | |
| Severe | 15 (19.5) | 453 (18.1) | 646 (14.7) | |

Abbreviations: PCC, post-COVID condition

**Supplementary Table 6.** Prevalence of post-COVID condition (PCC) among Quebec healthcare workers according to varying definitions and by participant characteristics

| | ≥4 weeks post-acute COVID-19 | | | | | | | ≥12 weeks post-acute COVID-19 | | | |
|---|---|---|---|---|---|---|---|---|---|---|---|
| | Total | PCC (any symp) | | | PCC[a] (≥1 severe symp) | | | Total | PCC (any symp) | | |
| | | N | L % | p | N | L % | p | N | N | L % | p |
| Overall | 6061 | 2836 | 46.8 | | 1080 | 17.8 | | 1783 | 680 | 38.1 | |
| **Age** (years) | | | | <.01 | | | <.01 | | | | <.01 |
| 18-29 | 1422 | 540 | 38.0 | | 207 | 14.6 | | 441 | 132 | 29.9 | |
| 30-39 | 1568 | 686 | 43.8 | | 247 | 15.8 | | 480 | 178 | 37.1 | |
| 40-49 | 1580 | 835 | 52.9 | | 343 | 21.7 | | 467 | 217 | 46.5 | |
| 50-59 | 1141 | 601 | 52.7 | | 227 | 19.9 | | 302 | 118 | 39.1 | |
| 60-80 | 350 | 174 | 49.7 | | 56 | 16.0 | | 93 | 35 | 37.6 | |
| **Sex** | | | | <.01 | | | <.01 | | | | 0.01 |
| Men | 1274 | 488 | 38.3 | | 153 | 12.0 | | 406 | 133 | 32.8 | |
| Women | 4787 | 2348 | 49.1 | | 927 | 19.4 | | 1377 | 547 | 39.7 | |
| **Race / ethnicity** | | | | <.01 | | | 0.01 | | | | 0.03 |
| White | 4794 | 2326 | 49.3 | | 880 | 18.4 | | 1382 | 552 | 39.9 | |
| Black | 490 | 159 | 32.5 | | 67 | 13.7 | | 150 | 40 | 26.7 | |
| Hispanic | 173 | 73 | 42.2 | | 34 | 19.7 | | 63 | 21 | 33.3 | |
| Arab | 184 | 62 | 33.7 | | 25 | 13.6 | | 64 | 21 | 32.8 | |
| Asiatic | 143 | 48 | 33.6 | | 17 | 11.9 | | 40 | 13 | 32.5 | |
| Other /NR | 277 | 132 | 47.7 | | 57 | 20.6 | | 84 | 33 | 39.3 | |
| **Profession** | | | | <.01 | | | <.01 | | | | <.01 |
| Physicians | 243 | 84 | 34.6 | | 22 | 9.1 | | 69 | 24 | 34.8 | |
| Nurses | 1138 | 569 | 50.0 | | 212 | 18.6 | | 321 | 125 | 38.9 | |
| Nurse assistants | 484 | 265 | 54.8 | | 120 | 24.8 | | 134 | 64 | 47.8 | |
| Patient healthcare assistants | 1566 | 754 | 48.2 | | 335 | 21.4 | | 369 | 147 | 39.8 | |
| Housekeeping | 202 | 94 | 46.5 | | 38 | 18.8 | | 45 | 15 | 33.3 | |
| Admin / Managers | 604 | 299 | 49.5 | | 109 | 18.1 | | 198 | 92 | 46.5 | |
| Psychosocial workers | 221 | 103 | 46.6 | | 30 | 13.6 | | 95 | 38 | 40.0 | |
| Other | 1603 | 668 | 41.7 | | 214 | 13.4 | | 552 | 175 | 31.7 | |
| **Vaccination status[b]** | | | | <.01 | | | 0.11 | | | | 0.23 |
| Unvaccinated | 5598 | 2664 | 47.2 | | 1009 | 18.0 | | 1569 | 610 | 38.9 | |
| 1 dose 0-13 days | 232 | 105 | 45.3 | | 39 | 16.8 | | 68 | 21 | 30.9 | |
| 1 dose ≥14 days | 218 | 84 | 38.5 | | 32 | 14.7 | | 141 | 49 | 34.8 | |
| 2 doses | 13 | 3 | 23.1 | | 0 | 0.0 | | 5 | 0 | 0.0 | |
| **COVID-19 Severity** | | | | <.01 | | | <.01 | | | | <.01 |
| Non-hospitalized | 5943 | 2746 | 46.2 | | 1033 | 17.4 | | 1746 | 653 | 37.4 | |
| Hospitalized | 118 | 90 | 76.3 | | 47 | 39.8 | | 37 | 27 | 73.0 | |

[a]For these definitions the 503 cases who were already recovered at the time of the survey but reporting symptoms lasting more than 4 weeks were excluded

[b]Vaccination status defined as: 1 dose administered 0-13 or ≥14 days before illness onset or 2 doses with the second dose administered at any moment before illness onset. Test comparing vaccinated with at least one dose 14 days before symptoms' onset with non-vaccinated or vaccinated with one dose days 0-13

Abbreviations: L%, line percentage; NR, no response; PCC, post-COVID condition; symp, symptom

**Supplementary Table 7.** Association of 4-week post-COVID condition with the duration of COVID-19 sick leave among Quebec healthcare workers with non-hospitalized and hospitalized COVID-19 (negative binomial regression model)

| | All COVID-19 cases[a] | | | | COVID-19 cases with finished sick leave[b] | | | |
|---|---|---|---|---|---|---|---|---|
| | % still on leave | Mean ± Std | Median | Adjusted mean ratio; 95% CI | % not fully recovered when returned to work | Mean ± Std | Media n | Adjusted mean ratio; 95% CI |
| **NON-HOSPITALIZED CASES** | | | | | | | | |
| N (denominator) | 5,302 | 5,302 | 5,302 | 5,302 | 4,999 | 4,999 | 4,999 | 4,999 |
| All non-hospitalized cases | 5.7 | 17.4 ± 17 | 13.0 | | 49.5 | 15.0 ± 12 | 12.0 | |
| **Post-COVID condition** | | | | | | | | |
| Non-PCC cases | 1.3 | 13.8 ± 12 | 12.0 | ref | 26.4 | 13.2 ± 11 | 11.0 | ref |
| PCC cases | 9.8 | 20.8 ± 20 | 14.0 | 1.5; 1.4-1.5 | 73.1 | 16.8 ± 13 | 13.0 | 1.3; 1.2-1.3 |
| **PCC by severity** | | | | | | | | |
| Non-PCC cases | 1.3 | 13.8 ± 12 | 12.0 | ref | 26.4 | 13.2 ± 11 | 11.0 | ref |
| Only mild symptoms | 1.8 | 16.6 ± 14 | 13.0 | 1.2; 1.1-1.3 | 62.9 | 15.8 ± 12 | 13.0 | 1.2; 1.1-1.2 |
| ≥1 moderate symptom | 7.6 | 19.7 ± 18 | 14.0 | 1.4; 1.3-1.5 | 74.3 | 16.8 ± 12 | 13.0 | 1.3; 1.2-1.3 |
| ≥1 severe symptom | 19.8 | 26.0 ± 25 | 16.0 | 1.8; 1.7-1.9 | 82.3 | 18.0 ± 15 | 14.0 | 1.3; 1.3-1.4 |
| **HOSPITALIZED CASES** | | | | | | | | |
| N (denominator) | 105 | 105 | 105 | 105 | 65 | 65 | 65 | 65 |
| All hospitalized cases | 38.1 | 41.1 ± 27 | 36.0 | | 63.1 | 28.9 ± 17 | 24.0 | |
| **Post-COVID condition** | | | | | | | | |
| Non-PCC cases | 11.1 | 21.2 ± 11 | 17.0 | ref | 37.5 | 19.4 ± 10 | 16.0 | ref |
| PCC cases | 43.1 | 45,3 ± 27 | 41.0 | 2.1; 1.6-2.7 | 71.4 | 32.0 ± 18 | 27.0 | 1.8; 1.4-2.4 |
| **PCC by severity** | | | | | | | | |
| Non-PCC cases | 11.1 | 21.2 ± 11 | 17.0 | ref | 37.5 | 19.4 ± 10 | 16.0 | ref |
| Only mild symptoms | 11.8 | 38.1 ± 15 | 39.0 | 1.8; 1.2-2.5 | 40.0 | 37.5 ± 16 | 39.0 | 2.0; 1.4-2.8 |
| ≥1 moderate symptom | 33.3 | 39.7 ± 28 | 35.0 | 1.8; 1.3-2.4 | 85.0 | 29.0 ± 22 | 21.0 | 1.6; 1.2-2.2 |
| ≥1 severe symptom | 65.0 | 52.5 ± 29 | 43.0 | 2.5; 1.9-3.3 | 85.7 | 30.5 ± 13 | 28.5 | 2.1; 1.5-2.9 |

[a] For those still in sick leave at the time of the survey, duration has been estimated as number of days from the beginning of the sick leave till the date of survey's participation

[b] Inverse probability weighting has been use to account for cases still on sick leave

Abbreviations: CI, confidence intervals; HCW, healthcare worker; PCC, Post-COVID condition; Ref, reference category; Std, standard deviation

Note: Negative binomial regression models adjusted for sex, age (5 categories: 18-29, 30-39, 40-49, 50-59, >60 years), race / ethnicity (6 categories: non-Hispanic White, Black, Hispanic, Arab, Asiatic, other) and occupation (5 categories: Physician, nurse, nurse assistant, patient healthcare assistant, housekeeping, administration /management, psychosocial workers, other).

**Supplementary Material 1.** Questionnaire for cases and controls

### Questionnaire for healthcare workers with confirmed COVID-19 (Cases)

(questions related to the post-COVID condition analysis)

#### SOCIO-DEMOGRAPHIC INFORMATION

1) Where do you live? Health region (1 to 18) [ _ ]
2) What is your mother tongue? ❒ French, ❒ English, ❒ Other
3) How old are you? [ _ ]
4) What is your gender? ❒ M ❒ F
5) Which of the following categories best describes you? ❒ Native (First Nations, Inuit, Métis), ❒ White, ❒ Asian, ❒ Black, ❒ Arab, ❒ Hispanic, ❒ Other, specify [ _ ] ❒ don't know, ❒ prefer not to answer

#### VACCINATION RECORD

6) Are you vaccinated against COVID-19? ❒ No ❒ Yes
   1st dose -> date [ _ ]
   2nd dose ->date [ _ ] ❒ I did not received a second dose

#### JOB DESCRIPTION

7) What is your primary job in the health care system?
   ❒ Security guard, ❒ Nursing Aide, ❒ Ambulance driver/paramedic, ❒ Volunteer, ❒ Stretcher Bearer, ❒ Cook or kitchen worker, ❒ Dentist, ❒ Special education teacher, ❒ Administrative/Managerial employee, ❒ Building maintenance employee, ❒ Housekeeping employee, ❒ Laundry service employee, ❒ Occupational Therapist, ❒ Student, intern or resident in any discipline, ❒ Dental hygienist, ❒ Nurse, ❒ Nursing Assistant, ❒ Respiratory therapist, ❒ Psychosocial worker, ❒ Physician, ❒ Nutritionist, ❒ Optometrist, ❒ Pharmacist, ❒ Physiotherapist, ❒ Patient healthcare assistant, ❒ Receptionist, ❒ Midwife, ❒ Laboratory technician, ❒ Pharmacy technician, ❒ Medical imaging technician (radiology, nuclear medicine, etc.) ❒ Other, specify: [ _ ]
8) How many years of experience do you have in this type of job? [ _ ] years, ❒ <1 year

#### DISEASE INFORMATION

9) According to your records from the Public Health survey in your area, **the date of onset of your first symptoms** is [ _ ] (yyyy/mm/dd).
   Would you like to confirm or correct the date listed above or enter the most accurate date if it was unknown in your file? (if you are not sure of the date, please enter an approximate date)
   [ _ ] (yyyy/mm/dd) ❒ I didn't have any symptoms

10) According to your records from the Public Health survey in your region, **the date of collection for the COVID-19 test is** [ _ ] (yyyy/mm/dd)
Would you like to confirm or correct the date listed above or enter the most accurate date if it was unknown in your file? [ _ ] (yyyy/mm/dd)

11) Have you had any symptoms of COVID-19? ❐ No, I had no symptoms ❐ Yes

12) Have you been hospitalized, including time in the emergency room, due to COVID-19 for more than 24 hours? ❐ No, ❐ Yes

13) Have you been admitted to the intensive care unit? ❐ No, ❐ Yes

14) On which date did you stop working due to COVID-19 disease? Date: [ _ ] (yyyy/mm/dd), ❐ I do not know, ❐ I did not have any sick leave

15) What was the date of your return to work (if you were on vacation, write the date when you felt capable of returning to work) Date: [ _ ] (yyyy/mm/dd), ❐ I do not know, ❐ I am still on sick leave

16) Did you feel fully recovered when returning to work? ❐ No, ❐ Yes, ❐ I do not know, ❐ Not applicable

17) How many weeks after the onset of your symptoms did you feel fully recovered? [ _ ] weeks, ❐ I still have symptoms of COVID-19.

18) If you still have symptoms, check all the symptoms that persist and their intensity:

| | |
|---|---|
| ❐ Fatigue: | ❐ Mild, ❐ Moderate, ❐ Severe |
| ❐ Fatigue level prior to COVID_19: | ❐ Mild, ❐ Moderate, ❐ Severe |
| ❐ Fever/sensation of fever: | ❐ Mild, ❐ Moderate, ❐ Severe |
| ❐ Cough: | ❐ Mild, ❐ Moderate, ❐ Severe |
| ❐ Shortness of breath: | ❐ Mild, ❐ Moderate, ❐ Severe |
| ❐ Wheezing: | ❐ Mild, ❐ Moderate, ❐ Severe |
| ❐ Chest pain: | ❐ Mild, ❐ Moderate, ❐ Severe |
| ❐ Sore throat: | ❐ Mild, ❐ Moderate, ❐ Severe |
| ❐ Headache: | ❐ Mild, ❐ Moderate, ❐ Severe |
| ❐ Muscle pain: | ❐ Mild, ❐ Moderate, ❐ Severe |
| ❐ Joint pain: | ❐ Mild, ❐ Moderate, ❐ Severe |
| ❐ Difficulty walking: | ❐ Mild, ❐ Moderate, ❐ Severe |
| ❐ Abdominal pain: | ❐ Mild, ❐ Moderate, ❐ Severe |
| ❐ Diarrhea: | ❐ Mild, ❐ Moderate, ❐ Severe |
| ❐ Loss of smell: | ❐ Mild, ❐ Moderate, ❐ Severe |
| ❐ Loss of taste: | ❐ Mild, ❐ Moderate, ❐ Severe |
| ❐ Other: specify _________: | ❐ Mild, ❐ Moderate, ❐ Severe |

### KESSLER PSYCHOLOGICAL DISTRESS SCALE (K6)

Over the last month, how often did you feel:

19) Nervous:
❒ Always, ❒ Often, ❒ Sometimes, ❒ Rarely, ❒ Never, ❒ I don’t know, ❒ I choose not to answer

20) Hopeless:
❒ Always, ❒ Often, ❒ Sometimes, ❒ Rarely, ❒ Never, ❒ I don’t know, ❒ I choose not to answer

21) Restless or fidgety:
❒ Always, ❒ Often, ❒ Sometimes, ❒ Rarely, ❒ Never, ❒ I don’t know, ❒ I choose not to answer

22) So depressed that nothing could cheer you up:
❒ Always, ❒ Often, ❒ Sometimes, ❒ Rarely, ❒ Never, ❒ I don’t know, ❒ I choose not to answer

23) That everything was an effort:
❒ Always, ❒ Often, ❒ Sometimes, ❒ Rarely, ❒ Never, ❒ I don’t know, ❒ I choose not to answer

24) Worthless:
❒ Always, ❒ Often, ❒ Sometimes, ❒ Rarely, ❒ Never, ❒ I don’t know, ❒ I choose not to answer

### COGNITIVE EVALUATION

BEFORE your COVID-19, how often did you (check the severity of the symptom):

25) Have difficulty to concentrate or maintain your attention at work or in your activities?
❒ Never, ❒ Rarely, ❒ Sometimes, ❒ Often, ❒ Very often

26) Have difficulty to organize your work or activities?
❒ Never, ❒ Rarely, ❒ Sometimes, ❒ Often, ❒ Very often

27) Forget things at work or in your activities?
❒ Never, ❒ Rarely, ❒ Sometimes, ❒ Often, ❒ Very often

28) Lose things that you need for your work or activities?
❒ Never, ❒ Rarely, ❒ Sometimes, ❒ Often, ❒ Very often

PRESENTLY, how often do you (check the severity of the symptom):

29) Have difficulty to concentrate or maintain your attention at work or in your activities?
❒ Never, ❒ Rarely, ❒ Sometimes, ❒ Often, ❒ Very often

30) Have difficulty to organize your work or activities?
❒ Never, ❒ Rarely, ❒ Sometimes, ❒ Often, ❒ Very often

31) Forget things at work or in your activities?
❒ Never, ❒ Rarely, ❒ Sometimes, ❒ Often, ❒ Very often

32) Lose things that you need for your work or activities?
❒ Never, ❒ Rarely, ❒ Sometimes, ❒ Often, ❒ Very often

**Questionnaire for healthcare workers without COVID-19 (Controls)**
(questions related to the post-COVID condition analysis)

### SOCIO-DEMOGRAPHIC INFORMATION

1) Where do you live? Health region (1 to 18) [ _ ]
2) What is your mother tongue? ❒ French, ❒ English, ❒ Other
3) How old are you? [ _ ]
4) What is your gender? ❒ M ❒ F
5) Which of the following categories best describes you? ❒ Native (First Nations, Inuit, Métis), ❒ White, ❒ Asian, ❒ Black, ❒ Arab, ❒ Hispanic, ❒ Other, specify [ _ ] ❒ don't know, ❒ prefer not to answer

### VACCINATION RECORD

6) Are you vaccinated against COVID-19? ❒ No ❒ Yes
   1st dose -> date [ _ ]
   2nd dose ->date [ _ ] ❒ I did not received a second dose

### JOB DESCRIPTION

7) What is your primary job in the health care system?
❒ Security guard, ❒ Nursing Aide, ❒ Ambulance driver/paramedic, ❒ Volunteer, ❒ Stretcher Bearer, ❒ Cook or kitchen worker, ❒ Dentist, ❒ Special education teacher, ❒ Administrative/Managerial employee, ❒ Building maintenance employee, ❒ Housekeeping employee, ❒ Laundry service employee, ❒ Occupational Therapist, ❒ Student, intern or resident in any discipline, ❒ Dental hygienist, ❒ Nurse, ❒ Nursing Assistant, ❒ Respiratory therapist, ❒ Psychosocial worker, ❒ Physician, ❒ Nutritionist, ❒ Optometrist, ❒ Pharmacist, ❒ Physiotherapist, ❒ Patient healthcare assistant, ❒ Receptionist, ❒ Midwife, ❒ Laboratory technician, ❒ Pharmacy technician, ❒ Medical imaging technician (radiology, nuclear medicine, etc.) ❒ Other, specify: [ _ ]
8) How many years of experience do you have in this type of job? [ _ ] years, ❒ <1 year

### KESSLER PSYCHOLOGICAL DISTRESS SCALE (K6)

Over the last month, how often did you feel:

9) Nervous:
   ❒ Always, ❒ Often, ❒ Sometimes, ❒ Rarely, ❒ Never, ❒ I don't know, ❒ I choose not to answer
10) Hopeless:
   ❒ Always, ❒ Often, ❒ Sometimes, ❒ Rarely, ❒ Never, ❒ I don't know, ❒ I choose not to answer
11) Restless or fidgety:

❒ Always, ❒ Often, ❒ Sometimes, ❒ Rarely, ❒ Never, ❒ I don't know, ❒ I choose not to answer

12) So depressed that nothing could cheer you up:
❒ Always, ❒ Often, ❒ Sometimes, ❒ Rarely, ❒ Never, ❒ I don't know, ❒ I choose not to answer

13) That everything was an effort:
❒ Always, ❒ Often, ❒ Sometimes, ❒ Rarely, ❒ Never, ❒ I don't know, ❒ I choose not to answer

14) Worthless:
❒ Always, ❒ Often, ❒ Sometimes, ❒ Rarely, ❒ Never, ❒ I don't know, ❒ I choose not to answer

**COGNITIVE EVALUATION**

PRESENTLY, how often do you (check the severity of the symptom):

15) Have difficulty to concentrate or maintain your attention at work or in your activities?
❒ Never, ❒ Rarely, ❒ Sometimes, ❒ Often, ❒ Very often

16) Have difficulty to organize your work or activities?
❒ Never, ❒ Rarely, ❒ Sometimes, ❒ Often, ❒ Very often

17) Forget things at work or in your activities?
❒ Never, ❒ Rarely, ❒ Sometimes, ❒ Often, ❒ Very often

18) Lose things that you need for your work or activities?
❒ Never, ❒ Rarely, ❒ Sometimes, ❒ Often, ❒ Very often

19) What is your present level of fatigue?
❒ Mild, ❒ Moderate, ❒ Severe
